# Multi-ancestry MHC-pQTL mapping reveals disease-linked HLA protein networks and shared genetic architecture

**DOI:** 10.64898/2026.09.08.26362390

**Authors:** Sarah Sarguroh, Sam Morris, Esther Ng, Guillaume Butler-Laporte, Ruth Nanjala, Alfred Pozarickij, Ling Yang, Liming Li, Junshi Chen, Pei Pei, Jun Lv, Canqing Yu, Dianjianyi Sun, Zhengming Chen, Iona Y. Millwood, Robin G. Walters, Alexander J. Mentzer, Yang Luo

**Author notes:** CKB Research Tracker number: 2019-0029.

## Abstract

The major histocompatibility complex (MHC) is one of the most polymorphic and disease-relevant regions of the human genome, yet the downstream effects of MHC variation on circulating protein networks across ancestries remain incompletely defined. Here, we conducted multi-ancestry protein quantitative trait locus (pQTL) mapping in the UK Biobank (n = 43,762) and China Kadoorie Biobank (n = 3,977) to characterise how genetic variation within the MHC region shape plasma protein abundance. Among 2,920 plasma proteins tested, we identified significant MHC associations (13 *cis* and 606 *trans*), with most signals showing consistent effects across ancestries, while also identifying an ancestry-enriched association with EDAR in East Asians. Local genetic correlation analyses identified distinct co-regulated protein networks associated with HLA class I and class II regions, consistent with their respective known immune functions. We also conducted HLA-specific colocalisation, and found multiple HLA *trans*-regulated proteins associations which colocalised with immune-mediated traits at the same classical HLA gene, including colocalisation between B2M and multiple sclerosis at HLA-A (PP = 0.991). Together, these findings describe the principal downstream proteomic consequences of MHC variation and provide a framework for linking disease-associated HLA genes to immune protein networks.

## Main

The plasma proteome sits at the interface between genetic variation and human physiology. Circulating proteins function as effectors, signalling molecules and structural components across nearly all biological processes. Their abundance is under substantial genetic control and can reflect tissue-level regulatory processes, either through active secretion, cellular leakage or shared genetic effects on expression across compartments. Protein quantitative trait loci (pQTLs), genetic variants which are associated with protein abundance, provide a powerful way to connect genomic variation to biological function and disease risk (1–3). The rapid growth of proteogenomic resources, including the UK Biobank Pharma Proteomics Project (UKB-PPP), which profiled over 2,900 proteins across more than 50,000 individuals has accelerated discovery and highlighted the value of population-scale proteomic studies (1).

The major histocompatibility complex (MHC) is particularly compelling for proteogenomic investigation. Spanning approximately 4 Mb on chromosome 6p21, it is the most gene-dense and disease-associated region of the human genome (4). Within the MHC, the classical human leukocyte antigen (HLA) genes encode antigen-presenting molecules that are central to adaptive immunity (5). Variation within the peptide-binding groove influences susceptibility to a wide range of immune-mediated diseases, including rheumatoid arthritis, multiple sclerosis, and type 1 diabetes, as well as drug hypersensitivity, transplant compatibility and infection outcomes (6,7). Previous pQTL studies have highlighted a large number of *trans*-pQTL signals in the MHC region (8), indicating that its regulatory effects extend far beyond local gene products. However, the MHC region has remained comparatively under-investigated. Many studies have excluded the MHC region entirely (9), treated it as a single locus (10,11), or focused only on *cis-*effects (12). Moreover, the limited genetic ancestral diversity in existing studies has left the generalisability of HLA-protein associations across populations largely unresolved (13,14).

Here, we present a multi-ancestry pQTL analysis of the MHC region by integrating data from the UK Biobank (UKB) and China Kadoorie Biobank (CKB). We aimed to: (1) perform a multi-ancestry MHC-wide analysis of both *cis-* and *trans*-pQTLs across 2,920 plasma proteins, including the identification of ancestry-enriched signals; (2) characterize the distal protein networks regulated by classical HLA class I and HLA class II variation; (3) determine the extent of shared genetic regulation among HLA-associated proteins; and (4) evaluate how these HLA-regulated proteomic effects relate to disease biology.

## Results

### Multi-ancestry analysis defines the landscape of MHC-regulated plasma proteins

We systematically identified genetic variants in the MHC region, including SNPs, classical HLA alleles and amino acid polymorphisms, that regulate circulating protein levels in the UK Biobank (UKB; *n* = 43,762) and the China Kadoorie Biobank (CKB; *n* = 3,977). We tested associations between 20,981 imputed variants and 2,920 plasma proteins measured using the Olink Explore platform. To minimise the risk of any imputation panel-specific biases, we imputed HLA variants using the same multi ancestry imputation panel for each cohort (15). To maximise discovery in this genetically complex region, we applied two complementary strategies. First, we performed a cohort-level meta-analysis that combined summary-level association results, and secondly, a joint analysis that modelled both cohorts in a unified framework using individual-level genotype and proteomic data (**Fig. 1**).

**Fig. 1.**
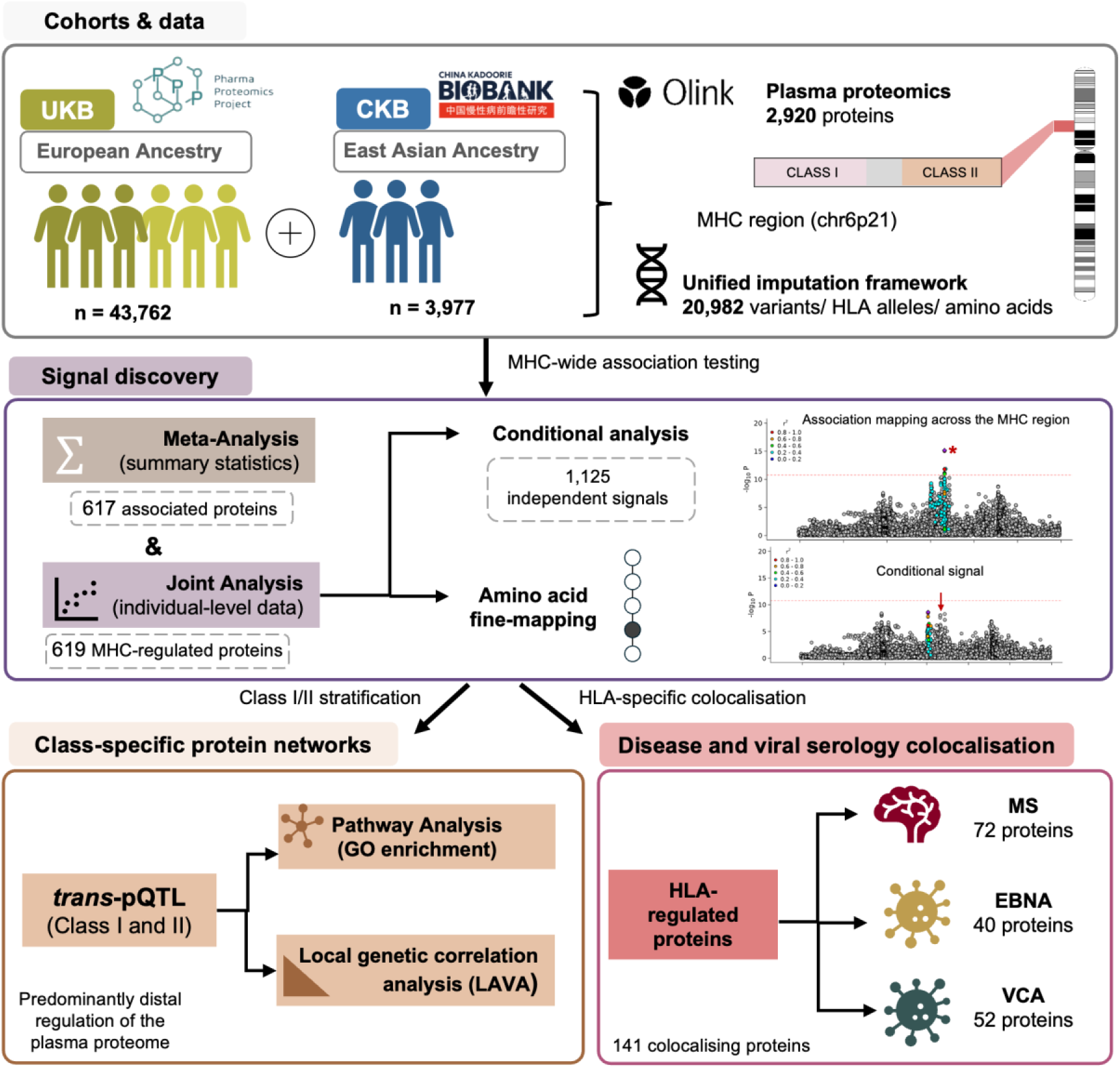
Overview of the multi-ancestry MHC-pQTL study design. Plasma proteomic data from UK Biobank participants of European ancestry and China Kadoorie Biobank participants of East Asian ancestry were analysed using a shared multi-ancestry HLA imputation framework across the MHC region. Multi-ancestry MHC-pQTL discovery was performed using both summary-statistic meta-analysis and joint individual-level analysis, followed by conditional analysis and amino-acid fine-mapping to resolve independent HLA signals. Significant MHC-regulated proteins were then used to define HLA class I- and class II-specific *trans*-pQTL networks, which were characterised using pathway enrichment and local genetic correlation analysis. Finally, HLA-specific colocalisation was used to identify MHC-regulated proteins sharing plausible causal HLA genes with multiple sclerosis and Epstein–Barr virus serological traits, including EBNA and VCA antibody responses.

In our separate cohort-specific analyses, we applied a stringent Bonferroni-adjusted threshold (P < 5 × 10⁻^8^/2,920 = 1.71 × 10⁻¹¹) and observed more proteins with at least one pQTL in UKB (n=603 proteins, 20.65% of total) than in CKB (n=72 proteins, 2.5% of total), consistent with the larger UKB sample size (**Fig. 2a**). When downsampling UKB to match the CKB sample size (n = 3,960), we saw comparable discovery rates across ancestries, with the number of proteins reaching significance in downsampled UKB similar to that observed in CKB (mean = 70.6 ± 0.7 across 10 iterations vs 72; **Supplementary Fig. 1**). We saw UKB effect sizes were highly concordant with those reported by Krishna et al. (8), who conducted a similar MHC-pQTL analysis in a slightly different subset of the same cohort (Spearman ρ = 0.9; **Supplementary Fig. 2**).

**Fig. 2.**
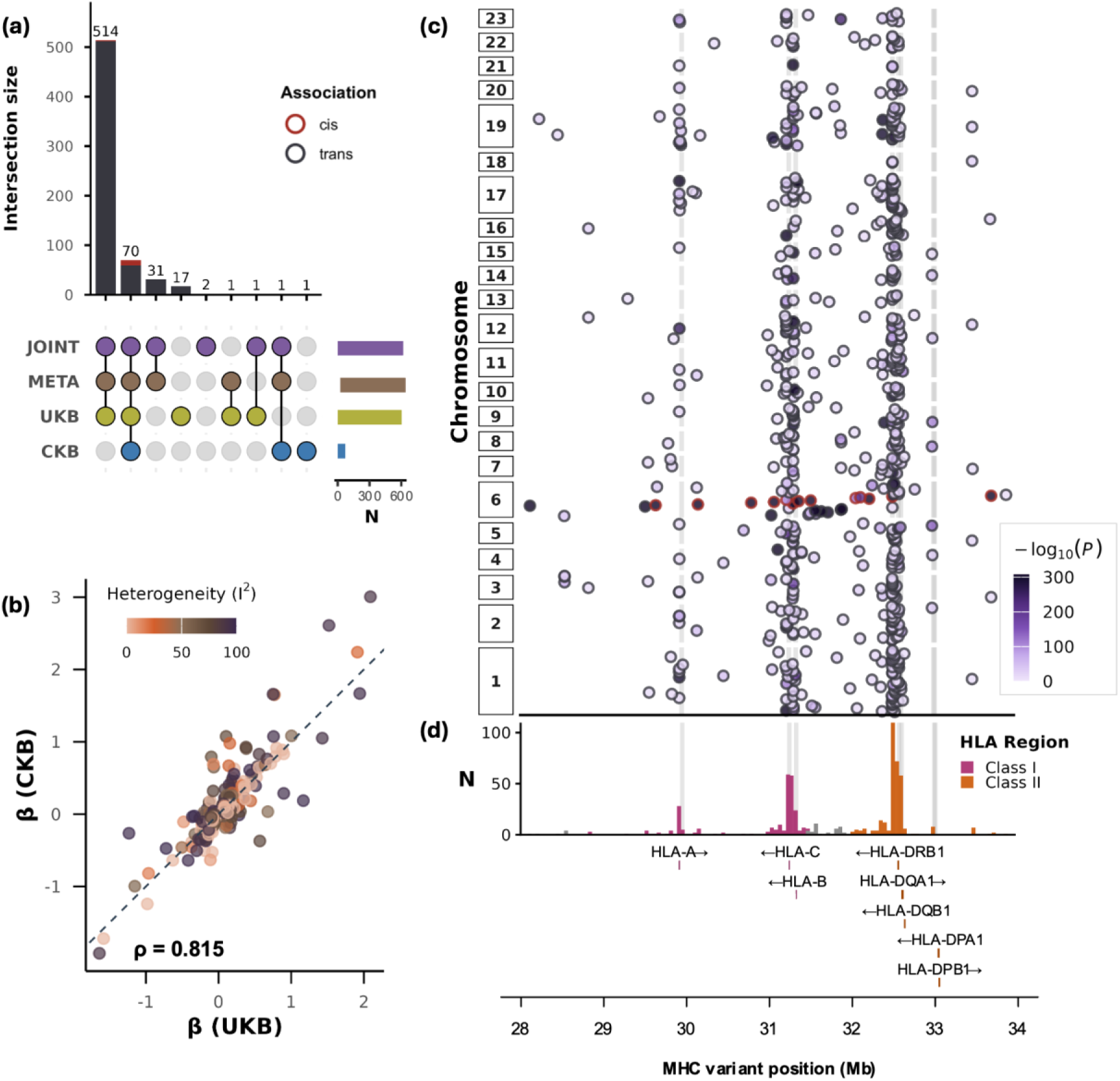
Multi-ancestry MHC-pQTL discovery and genomic distribution. **(a)** Overlap of genome-wide significant MHC-pQTL discoveries across the four analytical approaches: joint, fixed-effects meta-, UKB-only and CKB-only analysis. **(b)** Effect size concordance between UKB and CKB (Spearman ρ=0.815) for significant proteins in the meta-analysis, with points coloured by heterogeneity (I²). The dashed line indicates perfect concordance. **(c)** Genomic distribution of all genome-wide significant MHC-pQTL associations from the joint analysis. Each point represents a protein–variant association, coloured by −log₁₀P. The lower panel (**d**) shows the density of associations across the region.

We next performed a meta-analysis using cohort-specific summary statistics, which identified 617 proteins with at least one pQTL. We observed strong concordance in effect sizes between UKB and CKB pQTL signals (60.5% of signals have I² < 40%; Spearman ρ = 0.815, P = 5.74 × 10⁻²⁴⁸, **Fig. 2b**). These results indicate the genetic architecture of MHC-protein associations across individuals with European and East Asian genetic ancestries is largely similar.

Combining UKB and CKB in a joint pQTL association analysis not only increases power but also improves conditional analysis, as the two cohorts contribute differing linkage disequilibrium (LD) structures. After merging individual-level proteomic and genetic data and adjusting for both cohort effects and population structure, we identified 619 MHC-regulated proteins which largely overlapped with the 617 identified by meta-analysis (n=616 overlapping proteins, **Fig. 2a**). Given this concordance and the ability to perform conditional analysis directly on the joint model, we used it as the primary framework for all subsequent analyses. We performed iterative conditional analysis to identify proteins with multiple independent pQTLs and identified 1,125 independent pQTLs across 619 proteins, with 206 proteins (33.3%) exhibiting multiple independent signals (**Supplementary Fig. 3)**.

We next examined the regulatory architecture of MHC-pQTL associations in the joint analysis. Most of these associations were *trans*-acting (606/619, 98%), indicating that MHC variants act primarily through distal effects on the circulating proteome rather than through local *cis* regulation. In addition, these association signals were not uniformly distributed across the extended MHC region, but were strongly enriched within the eight imputed classical HLA genes (*HLA-A, B, C, DQA1, DQB1, DRB1, DPA1, DPB1*). Classical HLA alleles were also significantly over-represented among unique lead pQTL variants (OR = 4.80, Fisher’s exact P = 1.50 × 10⁻¹⁰), supporting a prominent role for antigen-presenting variation in MHC-mediated protein regulation (**Fig. 2c-d, Supplementary Fig. 4a-b**). Consistent with this interpretation, we also found MHC-regulated proteins were enriched among genes with tissue-specific expression in immune-related tissues such as lymph node, appendix, spleen and bone marrow (FDR < 0.05; **Supplementary Fig. 5**, **Supplementary Table 3**).

We further performed omnibus testing of HLA amino acid polymorphisms with significant MHC-signals, and identified multiple independent amino-acid positions associated with protein abundance. In total, we identified 690 amino acid-level signals across 503 proteins (P < 1.71 × 10⁻¹¹), with 133 proteins showing multiple independent signals (**Supplementary Fig. 6a**). These associated positions were enriched within the peptide-binding groove, which is defined by exon 2 and 3 for class I HLA genes and exon 2 for class II genes (OR = 2.83, Fisher’s exact P=3.1×10⁻⁵) (**Supplementary Fig. 6c**). This implies that variation in antigen presentation is a key mechanism linking HLA variation to downstream protein regulation. Notably, position 11 in HLA-DRB1 was the most frequently associated position, and was associated with 40 proteins, highlighting broad proteomic effects at a site with known functional importance in peptide binding and immune-mediated disease (16) (**Supplementary Fig. 6b**).

### HLA class I and class II variation shape distinct immune pathways and local protein regulatory networks

The marked enrichment of associations within classical HLA genes, together with their distinct LD structure across both populations (**Supplementary Fig. 7**), motivated us to dissect the distinct protein networks associated with HLA class I (*HLA-A, B, C*) and class II variation *(HLA-DRB1, DQA1, DQB1, DPA1, DPB1*; **Fig. 3a, b**). We focused on *trans*-acting associations because these are more likely to capture downstream immune regulatory mechanisms than local genetic effects on protein-coding genes within the MHC region.

**Fig. 3.**
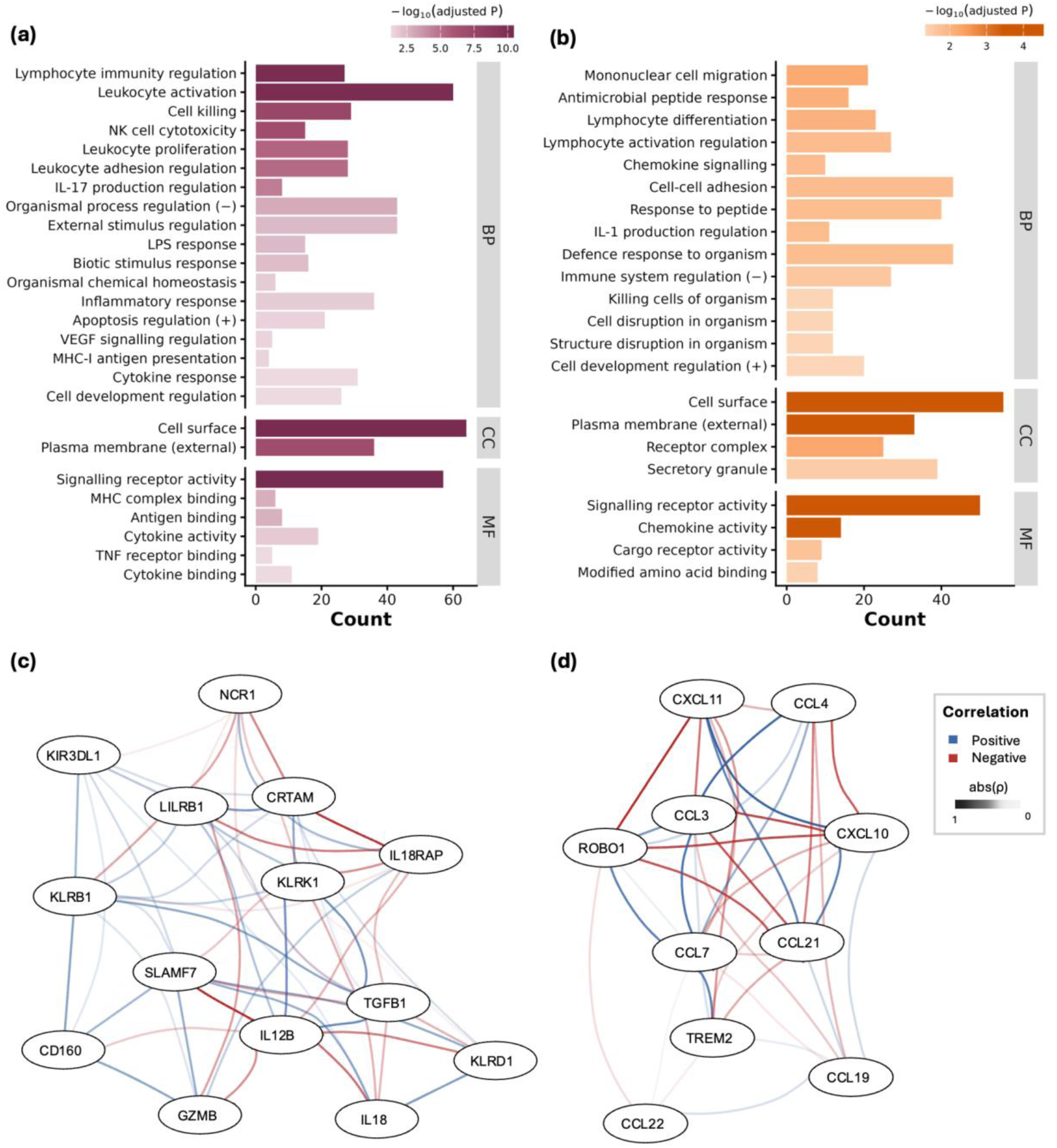
Functional enrichment and co-regulation networks of HLA class I- and class II-associated proteins. Gene Ontology enrichment analysis of proteins with *trans*-pQTL associations mapped to (**a**) HLA class I genes or (**b**) class II genes. Bars show the number of proteins contributing to each enriched Gene Ontology term, grouped by biological process (BP), cellular component (CC) and molecular function (MF) and coloured by −log₁₀(adjusted P). Local genetic correlation networks among proteins contributing to **(c)** NK cell-mediated cytotoxicity (class I) and **(d)** chemokine signalling (class II), estimated using LAVA within the class I (chr6:29.5–31.5 Mb) and class II (chr6:32.0–33.5Mb) loci respectively. Nodes represent proteins and edges indicate significant pairwise local genetic correlations (P < 1.55 × 10 ⁻⁶). Edge colour indicates the direction of correlation (blue, positive; red, negative) and edge opacity is proportional to |ρ|. Full pairwise correlation results are reported in **Supplementary Table 9-10**.

In total, we identified 172 proteins with class I-specific *trans* associations and 195 with class II-specific *trans* associations. These two groups showed distinct functional enrichments; class I-associated proteins were enriched for cytotoxic immune responses and inflammatory effector mechanisms (**Fig. 3a**), whereas class II-associated proteins were preferentially enriched for chemokine signalling, leukocyte migration and T cell communication pathways (**Fig. 3b, Supplementary Table 7-8).** These contrasting profiles mirrored the established biology of HLA class I and class II molecules and show that genetic variation in the two antigen-presentation systems shapes distinct downstream immune mechanisms (6,8).

We next investigated how proteins within each class-specific network were genetically correlated with one another. To investigate the local genetic correlation (r_g_) between proteins, we applied LAVA (17), a tool for robust local r_g_ analysis, to pairs of distal proteins both regulated either by HLA class I or class II variations. Following univariate filtering for significant local heritability, bivariate tests were performed on all eligible protein pairs within each class, yielding 14,706 tests in class I and 17,578 in class II. After Bonferroni correction across both classes (P < 0.05/32,284 = 1.55 × 10⁻⁶), 26,390 pairs of proteins showed significant r_g_, representing approximately 81.7% of all tested correlations, reflecting the extensive shared genetic architecture within the MHC (**Supplementary Fig 8-9**).

Among these, 3,459 pairs showed particularly strong genetic correlation (|ρ| ≥ 0.7), indicating dense locus-specific co-regulation within the MHC region (**Supplementary Table 9-10**). To illustrate the structure of these networks, we visualised local genetic correlations among proteins contributing to representative class-specific pathways: NK cell-mediated cytotoxicity for class I (**Fig. 3c**) and chemokine signalling for class II (**Fig. 3d**).

To further examine the robustness of our class-specific LAVA analyses, we performed a permutation test to assess whether the observed within-class enrichment was specific to the predefined HLA class assignments rather than arising from random assignment of proteins to classes. Class labels were randomly reassigned among the 172 class I- and 195 class II-significantly associated proteins while preserving the original group sizes. Across 1,000 permutations, the observed number of significant class I–class I pairs at the class I locus exceeded the permutation distribution (permutation P < 0.001), while the corresponding number of significant class II–class II pairs at the class II locus was also greater than expected under random assignment (permutation P = 0.002; **Supplementary Fig. 10**).

### HLA colocalisation links HLA-regulated proteins to MS and EBV serological responses

Having defined HLA-regulated protein networks, we next investigated whether these proteins could provide plausible molecular insights into how HLA variants might regulate disease risk and immune response. Here, we focused on multiple sclerosis (MS) and Epstein–Barr virus (EBV) serology, including Epstein-Barr nuclear antigen (EBNA) and viral capsid antigen (VCA) antibody responses.

We applied HLA-colocalisation, a Bayesian summary-statistic-based method to test whether two traits share the same gene-level HLA association pattern rather than being associated with just the same individual variant (18). Colocalisation provides evidence that two traits share a common causal variant at a locus. This gene-level approach is well suited to the complex LD and multi-allelic architecture of the HLA genes.

Using a posterior colocalisation probability threshold of > 0.90, we identified 72 proteins which colocalised with the MS signal (19), 40 with EBNA serology, and 52 with VCA serology (20), yielding 141 unique proteins across all three endpoints (**Fig. 4a**). These colocalisations mapped to four HLA genes (HLA-A, HLA-C, HLA-DRB1 and HLA-DQB1) and were broadly aligned with their established immunological roles (**Supplementary Table 12**). Notably, VCA yielded more colocalised proteins than EBNA, particularly at HLA-DRB1. As VCA and EBNA reflect lytic and latent phases of EBV biology, respectively, this pattern suggests that HLA control of antibody responses to lytic EBV antigens may engage a broader downstream circulating protein network than responses to the latent antigen EBNA. This interpretation is consistent with prior UKB analyses showing a positive genetic correlation between VCA, but not EBNA, and MS (18), and with recent evidence that CD4+ T cells in MS patients preferentially target EBV late lytic antigens rather than latent antigens (21).

**Figure 4.**
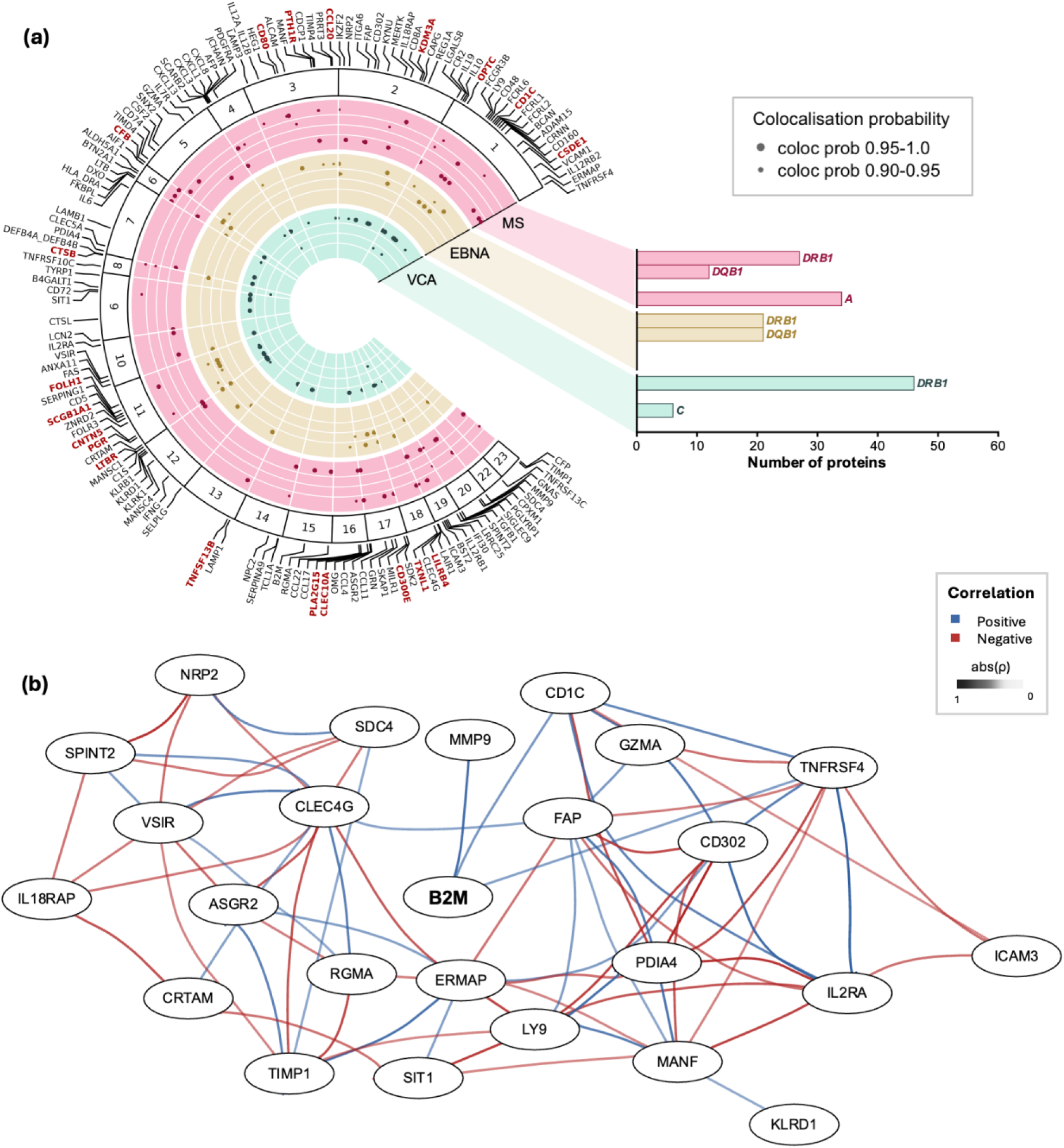
HLA colocalisation links HLA-regulated proteins to MS and EBV serological responses. **(a)** HLA-regulated proteins with posterior colocalisation probability > 0.90 with multiple sclerosis (MS), viral capsid antigen (VCA) antibody response and Epstein-Barr nuclear antigen (EBNA) antibody response, arranged by chromosomal location of the protein-coding gene. Points indicate colocalising protein–trait pairs, with larger points denoting posterior colocalisation probability >0.95. Proteins that are colocalised with more than one trait are highlighted in red. The bar plot summarises the number of colocalising proteins per trait at each HLA gene. Full results are reported in **Supplementary Table 12**. **(b)** Local genetic correlation network of HLA-A-regulated proteins colocalising with MS. Nodes represent proteins and edges indicate significant local genetic correlations (|ρ| ≥ 0.7, P < 1.55 × 10⁻⁶). Blue and red edges indicate positive and negative correlations, respectively.

Among these signals, Beta-2-microglobulin (B2M) provides a particularly compelling example linking HLA-regulated protein variations to MS biology. We observed a strong colocalisation signal between B2M and MS (posterior probability = 0.991) at HLA-A, led by HLA-A*02:01, the strongest protective class I allele for MS (22). B2M is an essential structural partner of HLA class I molecules (23). Cerebrospinal fluid B2M levels have been reported to be elevated in MS patients relative to controls (24). In experimental autoimmune encephalomyelitis, B2m-deficient mice exhibit reduced spinal cord lesion burden (25), further supporting its role in MS-relevant immune activation.

More broadly, we identified 34 HLA-A-regulated proteins that colocalised with MS GWAS signals, predominantly driven by HLA-A*02:01 (**Supplementary Fig. 13**). Local genetic correlation analysis revealed both positive and negative co-regulation among these proteins within the class I region (**Fig. 4b**), indicating that HLA-A variation coordinates a diverse downstream immune programme rather than acting through a single effector molecule.

The co-regulated network included proteins involved in antigen presentation (B2M, CD1C), cytotoxic effector function (CRTAM, GZMA) and lymphocyte costimulation (IL2RA), several of which have prior support for MS relevance through genetic association, biomarker studies, or experimental models (26–29). Together, these proteins nominate an HLA-A-regulated protein network linking antigen presentation, immune activation, and leukocyte effector functions to MS-associated genetic variation.

### An ancestry-enriched HLA-DQB1 association with circulating EDAR levels

Across all tested proteins, EDAR reached the study-wide significance threshold in CKB but not in UKB (**Fig. 5a**). The lead signal was HLA-DQB1*05:02, a class II allele with established roles in peptide antigen presentation and prior associations with autoimmune diseases (30,31). Each copy of DQB1*05:02 was associated with higher circulating EDAR levels in CKB (β = 0.279, SE = 0.040, P = 3.70×10⁻¹²), whereas no association was observed in UKB (β = 0.017, SE = 0.038, P = 0.655) (**Fig. 5b**). In the joint analysis, the combined signal did not reach the study-wide significance threshold (β = 0.141, SE = 0.028, P = 3.65×10⁻⁷), consistent with an ancestry-enriched effect that is diluted when pooled with the larger European sample. HLA-DQB1*05:02 was approximately 10x more frequent in CKB than in UKB (MAF = 0.079 vs 0.0078), providing greater power to detect its association in the East Asian cohort.

**Figure 5.**
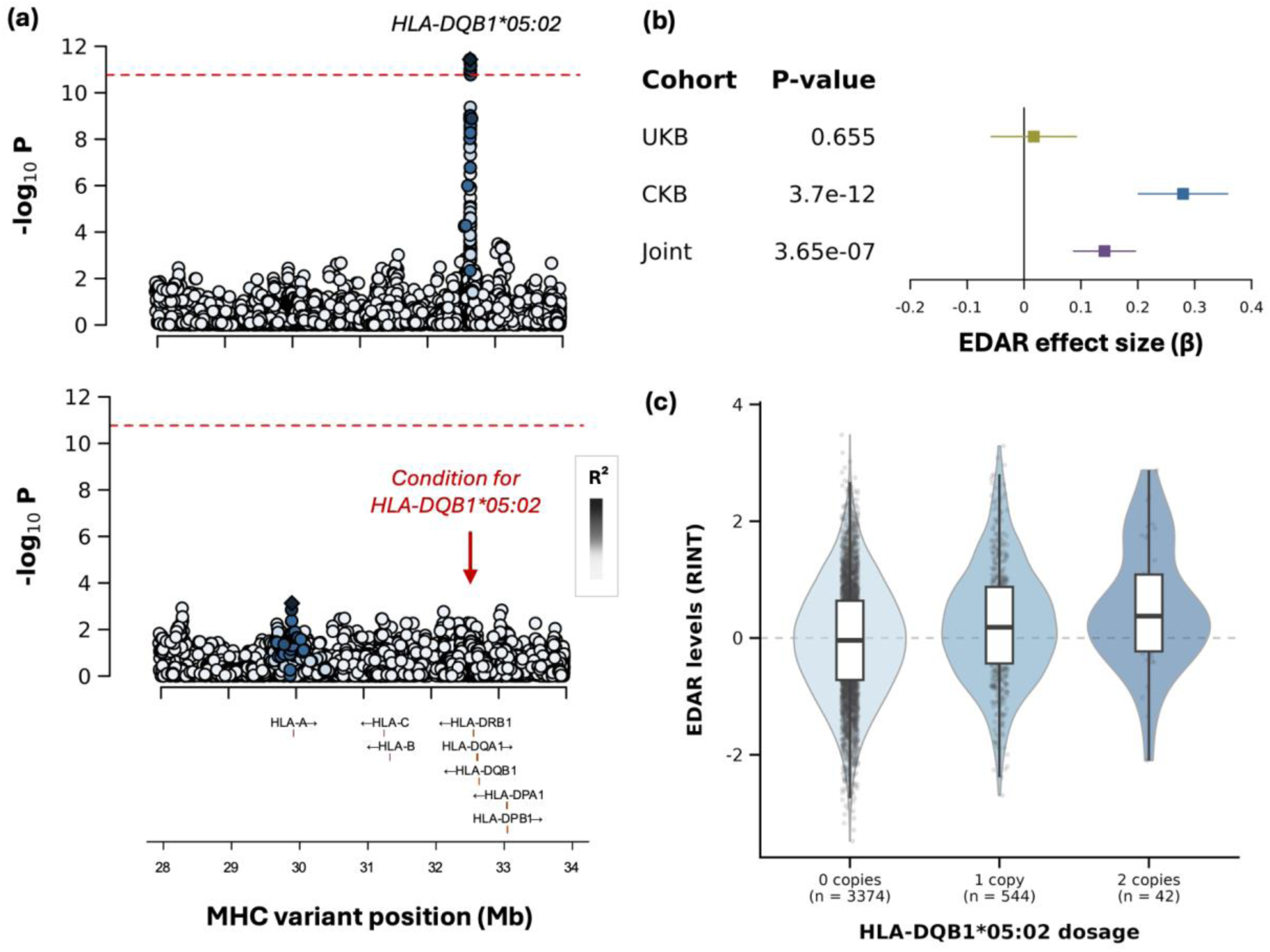
An ancestry-enriched HLA-DQB1 association with circulating EDAR levels. **(a)** Association plots for EDAR across the extended MHC region in CKB. The upper panel shows the CKB-only analysis, with points coloured by LD (R²) with the lead variant HLA-DQB1*05:02. The dashed red line indicates the study-wide significance threshold (P < 1.71 × 10⁻¹¹). The lower panel shows the conditional association analysis after adjustment for HLA-DQB1*05:02, showing attenuation of the primary signal and no additional independent association reaching the study-wide threshold. **(b)** Effect estimates for HLA-DQB1*05:02 on circulating EDAR levels in UKB, CKB and the joint analysis. Horizontal lines indicate 95% confidence intervals. **(c)** Distribution of EDAR protein levels (RINT) by HLA-DQB1*05:02 dosage in CKB participants.

To assess whether the result could be explained by differences in protein pre-processing, we compared the EDAR distributions across two cohorts. Raw EDAR NPX values showed lower variance in CKB than in UKB, but the two distributions aligned closely following residualisation and inverse-normal transformation **(Supplementary Fig. 19)**, indicating that the association was not driven by cohort differences in the marginal protein distribution.

*EDAR* encodes Ectodysplasin A Receptor, a member of the tumour necrosis factor (TNF) receptor family that is essential for the development of ectoderm-derived tissues, including hair follicles, teeth and sweat glands (31,32), through activation of NF-κB signalling in response to its ligand ectodysplasin A (EDA). Interestingly, EDAR also harbours a well-characterised signal of recent positive selection in humans, which enhances NF-κB signalling and has risen to high frequency in East Asian populations (33,34). Together, these observations underscore how multi-ancestry HLA-pQTL mapping can surface biologically meaningful candidates that European-only studies would miss.

## Discussion

In this study, we provide a large-scale, multi-ancestry characterisation of the effects of MHC genetic variation on the circulating proteome. By integrating plasma proteomic and HLA-region genetic data from individuals of European and East Asian ancestry, we identified 619 proteins regulated by variation within the MHC, the great majority of which were affected in *trans*. The effects of significant associations were broadly concordant between UKB and CKB, indicating that much of the downstream proteomic architecture of the MHC is shared across these populations. At the same time, the inclusion of CKB enabled the identification of an ancestry-enriched association between HLA-DQB1*05:02 and circulating EDAR levels that would not have been detected in the substantially larger European-ancestry cohort. By combining conditional association analysis, amino-acid-level mapping, pathway enrichment, local genetic correlation and HLA-specific colocalisation, our results extend previous studies of HLA-mediated protein regulation and provide a framework for connecting disease-associated HLA variation to downstream immune protein networks, while also revealing ancestry-enriched signals such as EDAR (8).

Several findings support antigen presentation as a major mechanism through which MHC variation influences circulating protein abundance. First, significant pQTLs were strongly enriched within the classical HLA genes, despite these genes occupying only a limited proportion of the extended MHC region. Second, most associations were *trans-*acting, indicating that the principal consequences of HLA variation are not restricted to the expression of locally encoded proteins. Third, amino-acid associations were enriched within the peptide-binding regions of HLA molecules, with HLA-DRB1 position 11 influencing particularly large numbers of proteins. Together, these results suggest that differences in the peptide repertoires presented by HLA molecules may alter the activation, differentiation or abundance of immune-cell populations, thereby generating broad downstream changes in circulating proteins. Nevertheless, peptide-binding enrichment does not exclude additional mechanisms, including effects on HLA stability, cell-surface expression, receptor engagement or cell-intrinsic signalling in antigen-presenting cells (8,23).

The separation of class I- and class II-regulated protein sets further showed that the two antigen-presentation systems influence distinguishable, although interconnected, immune programmes. Class I-associated proteins were enriched for NK-cell cytotoxicity, leukocyte activation and inflammatory effector functions, consistent with the roles of HLA class I molecules in CD8⁺ T-cell and NK-cell biology (6,35). In contrast, class II-associated proteins were preferentially enriched for chemokine signalling, leukocyte migration and intercellular immune communication, compatible with the central role of class II molecules in CD4 ⁺ T-cell activation and antigen-presenting-cell function (6,36). These results extend the interpretation of individual HLA-pQTL associations by showing that class I and class II variation can be organised into coherent downstream proteomic programmes rather than collections of unrelated protein associations.

Local genetic correlation analysis provided additional evidence that these proteins form structured locus-specific networks. More than 80% of the tested protein pairs showed significant local genetic correlation, and many exhibited strong positive or negative correlations within the class I or class II regions. Positive correlations may represent proteins influenced in the same direction by shared HLA alleles, whereas negative correlations could arise when the same alleles promote one immune programme while suppressing another. Such patterns are compatible with coordinated changes in immune-cell activation, differentiation or trafficking. However, local genetic correlation should not be interpreted as evidence that two proteins interact physically or that one regulates the other. In the MHC, correlations may also reflect extended LD, multiple correlated HLA alleles or distinct causal residues that are incompletely resolved statistically. The networks therefore provide a representation of shared local genetic architecture and a basis for prioritising protein groups for functional investigation, rather than definitive molecular pathways.

Using an HLA-specific colocalisation approach, we also showed that the HLA-regulated proteins identified could be candidate biomarkers underlying HLA-disease associations, providing a basis for the functional work needed to explore their roles in diseases. For example, we showed that in MS, the protective HLA-A signal colocalised with B2M abundance, consistent with the close biological relationship between HLA-A and its invariant class I partner B2M. More broadly, the MS and EBV results support the idea that HLA variation influences disease through coordinated protein networks rather than single downstream targets. The greater number of proteins colocalising with VCA than EBNA also suggests that HLA-regulated proteins may be particularly relevant to immune responses against active EBV infection.

A strength of this study is the use of a unified HLA imputation framework in both cohorts, reducing the likelihood that apparent cross-population differences arose solely from the use of different reference panels. The parallel use of cohort-specific meta-analysis and joint individual-level analysis also provided complementary evidence for association while enabling conditional analyses in the combined sample. In addition, the combination of variant-level association testing with HLA amino-acid mapping, reciprocal conditioning of class I and class II alleles, local genetic correlation and an HLA-specific colocalisation method allowed us to examine MHC regulation at several levels. This is particularly important in the HLA region, where conventional approaches developed for approximately independent biallelic variants may not adequately account for multi-allelic variation and long-range LD (37).

Our study also has several limitations. First, CKB is substantially smaller than UKB, which limits the power to detect ancestry-enriched signals. Second, imputation accuracy varies among HLA alleles and amino-acid residues, particularly for rare variation, and statistical conditioning cannot completely resolve causal alleles in the presence of extensive LD. Third, HLA colocalisation identifies shared genetic signals at a gene-level but does not establish whether a given protein mediates disease risk or reflects a downstream consequence of HLA variation. Finally, plasma protein measurements capture systemic biology and may miss tissue-specific immune processes.

As proteomics datasets continue to grow in both scale and ancestral diversity, the multi-ancestry framework applied here will be particularly valuable for resolving HLA-pQTL architecture and for identifying molecular links between MHC variation and immune-mediated disease.

## Methods

### Study Populations

This study uses proteogenomic data from two large prospective cohorts. The UK Biobank recruited approximately 500,000 adults aged 40–69 years from across the United Kingdom between 2006 and 2010 (1). For this study, we focused on the subset of participants from the UK Biobank Pharma Proteomics Project with available Olink Explore proteomics data at baseline (n = 53,018). We restricted the analysis to participants of genetically European ancestry (n = 43,762; Data Field ID = 22006).

The China Kadoorie Biobank recruited approximately 512,000 adults aged 30–79 years from five urban and five rural regions of China between 2004 and 2008 (38). We analysed 3,977 CKB participants of East Asian ancestry with available genotype and plasma proteomic data. Participant characteristics for both cohorts are summarised in **Supplementary Table 1.** Full details of the CKB genetic data, including genotyping array design can be found in Walters et al (39).

### HLA imputation and quality control

For each cohort, we separately imputed variants within the MHC region using the Michigan Imputation Server with the Four-digit Multi-ancestry HLA reference panel v2 (15). Imputation was performed using the GRCh38/hg38 genome build. A unified variant set was constructed by merging the imputed genotype data from both cohorts, retaining variants with MAF>0.1% and genotype missingness <90% across the combined sample, yielding 20,981 variants (18,115 SNPs, 2,585 amino acid polymorphisms and 281 classical HLA alleles). Given the known effects of balancing selection and long-range linkage disequilibrium on allele frequency distributions within the MHC region (37), we did not apply Hardy–Weinberg equilibrium filtering. This unified variant set was used for all downstream analyses.

### Plasma protein measurements and quality control

Plasma protein levels were measured using the Olink Explore platform (1), with 2,923 and 2,930 annotated proteins measured in UKB and CKB, respectively, as normalised protein expression (NPX) values on a log₂ scale. Protein processing followed the workflow described in the UKB-PPP flagship paper (1). Following harmonisation of the Olink assay measurements across cohorts, 2,921 proteins were retained in both datasets. One protein (GLIPR1) was excluded due to >90% missingness in UKB and was additionally removed from CKB to maintain a consistent protein set, yielding 2,920 proteins retained for analysis.

Sample-level outliers were identified using two complementary approaches applied within each cohort. First, for each individual, the median and interquartile range (IQR) of NPX values across all proteins were computed; individuals whose per-sample median or IQR exceeded ± 5 standard deviations from the cohort mean were flagged (UKB: 158; CKB: 15). Second, remaining missing values were imputed with the within-cohort column mean, consistent with the UKB-PPP flagship analysis (1), and PCA was performed on the imputed proteomic matrix; individuals with PC1 or PC2 scores exceeding ± 5 standard deviations from the mean were flagged (UKB: 22; CKB: 6). The union of outliers from both methods was removed (UKB: 180; CKB: 17; **Supplementary Fig. 17**).

Following outlier removal, protein levels were residualised within each cohort. In UKB, NPX values were residualised for sex, age, age², age×sex, age²×sex, recruitment centre, days to processing, protein batch, genotype batch, ten genetic PCs, and the top twenty proteomic PCs. In CKB, NPX values were residualised for sex, age, age², age×sex, age²×sex, recruitment region, genotyping array type, eleven genetic PCs (39), and the top twenty proteomic PCs. Residualised values were then subjected to rank-based inverse normal transformation (RINT) within each cohort.

The final analytical sample comprised 43,582 UKB and 3,960 CKB individuals across 2,920 proteins and 20,981 HLA-region variants. A summary of sample and protein QC steps is provided in **Supplementary Fig. 18**, and cohort characteristics are reported in **Supplementary Table 1**. Pre- and post-normalisation protein distributions for representative proteins are shown in **Supplementary Fig. 19.**

### Association testing and meta-analysis

HLA-pQTL mapping was performed using an additive linear regression model implemented in PLINK2 (40). As described above, all demographic, technical and proteomic covariates were regressed from protein levels within each cohort prior to RINT, so no additional covariates were included in the cohort-specific association models. For the joint analysis, individual-level genotype and protein data from both cohorts were combined. Since protein levels were residualised and normalised within each cohort independently, a binary cohort indicator (0 = UKB, 1 = CKB) was included as the sole covariate to account for any remaining systematic differences between cohorts.

Fixed-effects inverse-variance weighted meta-analysis was performed using PLINK2 (40) across 2,920 proteins with harmonised summary statistics available in both cohorts.

Across all four discovery models (UKB-only, CKB-only, fixed-effects meta-analysis and joint analysis), a protein was considered a protein was considered MHC-regulated if at least one variant–protein association reached Bonferroni-corrected threshold of P < 1.71 × 10⁻¹¹ (5 × 10⁻⁸/2,920). For each protein, the lead variant–protein pair was defined as the variant with the smallest P-value.

Associations were classified as cis if the protein-coding gene was located within the extended MHC region (chr6:28–34 Mb), and trans otherwise.

### Downsampling analysis for matched ancestry comparison

To assess whether discovery rates were comparable across ancestries at matched sample sizes, we downsampled UKB to match the CKB sample size (n = 3,960). Ten independent random subsets were drawn from the UKB participants, and MHC-wide pQTL analysis was performed on each using the same analytical pipeline and significance threshold (P < 1.71 × 10⁻¹¹).

### Conditional analysis

To identify independent association signals beyond the primary lead variant, we performed a stepwise conditional analysis for each genome-wide significant protein in the UKB-only, CKB-only, and the joint analysis. For each protein and dataset, association testing was repeated using PLINK2 (40) across all post-QC variants with the lead HLA-pQTL included as an additional covariate. The most significant variant remaining after conditioning was then evaluated against the genome-wide significance threshold (P < 1.71 × 10⁻¹¹); added to the independent set if significant, and the procedure repeated until no further variant reached significance. When multiple variants shared the same minimum P-value, we selected the variant with the largest absolute Z-score.

### Omnibus test

To resolve HLA-pQTL signals to the level of individual amino acid positions, we applied an omnibus test to all 619 proteins reaching genome-wide significance in the joint analysis, using the HLA-TAPAS framework (15). The omnibus test jointly evaluates whether the distinct amino acid residues at a given position are associated with protein levels, using an F-test with m−1 degrees of freedom, where m is the number of residues at the position. Analyses were performed separately for each protein, yielding omnibus P-values and per-haplotype effect estimates for each tested position. For each protein, the lead amino acid position was defined as the position with the smallest omnibus P-value.

Following identification of the lead amino acid position (the position with the smallest omnibus P-value), we performed stepwise conditional analysis to identify additional independent signals. At each step, the lead position was included as a covariate and all remaining positions within the same gene were re-tested. For proteins whose lead position mapped to HLA-DRB1, -DQA1, or -DQB1, all positions across these three genes were tested jointly given the extensive linkage disequilibrium across this superlocus. We limited the conditional analysis to a maximum of three independent amino acid positions per protein, and required each additional signal to reach the study-wide significance threshold (P < 1.71 × 10⁻¹¹).

To assess whether HLA-pQTL signals were enriched at residues in the peptide-binding groove, we compared lead positions with all polymorphic amino acid positions and were tested across the eight classical HLA genes. For HLA class I genes, peptide-binding site positions were defined by exons 2 and 3, and for class II genes by exon 2. Enrichment of lead omnibus positions within these regions was assessed using a one-sided Fisher’s exact test.

### Tissue enrichment

To examine tissue expression patterns, we used gene sets from the Human Protein Atlas (HPA), as reported by Uhlén et al (41), following the approach of Krishna et al (8). Genes were classified as tissue-specific if they were designated as ’tissue enriched’ (mRNA expression at least five-fold higher in one tissue compared to all other tissues) or ’tissue enhanced’ (mRNA expression at least five-fold higher than the average across all tissues) in the Human Protein Atlas. Enrichment of MHC-regulated proteins within each of 32 tissue gene sets was assessed using Fisher’s exact test, with all 2,920 Olink panel proteins as the background set. Significance was defined at FDR < 0.05 after Benjamini–Hochberg correction across tissues.

### Pathway enrichment

To dissect the contributions of HLA class I and class II genetic variation to plasma protein levels, we performed reciprocal conditioning analyses. Proteins were initially assigned to class I (n = 235) or class II (n = 312) based on the genomic position of their lead trans-pQTL variant within the HLA class I (chr6:29,500,000–31,500,000) or class II (chr6:32,000,001–33,500,000) sub-regions; 61 proteins with lead variants outside both sub-regions were excluded from this analysis. For proteins with a primary HLA class I association, we re-tested the association after conditioning on all classical HLA class II two-field alleles to isolate class I-specific effects, and vice versa for proteins with a primary class II association. Proteins that remained genome-wide significant after conditioning were assigned to class I-specific (172/235) or class II-specific (195/312) sets.

Gene ontology (GO) enrichment analyses were performed separately for each class-specific protein set using the *clusterProfiler* package (42) in R, with all proteins measured in both cohorts used as the background. Enrichment of GO terms (Biological Process, Cellular Components, and Molecular Functions) was tested using a hypergeometric test, and multiple testing correction was applied using the Benjamini–Hochberg false discovery rate (FDR < 0.05). Redundant GO terms were removed using semantic similarity-based simplification (simplify cutoff = 0.5) to retain representative terms (43).

### Local genetic correlation analysis

To characterise the shared local genetic architecture among HLA-pQTL signals, we applied Local Analysis of [co]Variant Association (LAVA) to summary statistics from the joint analysis (17). LAVA estimates bivariate local genetic correlations between pairs of traits within defined genomic loci while accounting for complex LD structure. An in-sample LD reference was derived from the joint analysis genotypes across the MHC region (MAF > 0.5%).

Analyses were restricted to the class I-specific and class II-specific trans-regulated protein sets defined by the reciprocal conditioning analysis described above. Two loci were defined to capture the canonical HLA class I and class II sub-regions: a class I locus (chr6:29,500,000– 31,500,000) and a class II locus (chr6:32,000,001–33,500,000). For each protein, local SNP heritability was first estimated within each locus. Proteins reaching a Bonferroni-corrected univariate significance threshold (class I: P < 2.9 × 10⁻⁴, 0.05/172; class II: P < 2.56 × 10⁻⁴, 0.05/195) were carried forward for bivariate testing.

Pairwise bivariate local genetic correlations were then estimated among all eligible proteins within each locus, yielding 14,706 tests in class I and 17,578 in class II, with significance assessed using a Bonferroni correction across all pairwise tests in both loci combined (0.05/32,284 = P < 1.55 × 10⁻⁶).

To assess the robustness of the observed within-class enrichment of significant local genetic correlations, we performed a permutation analysis in which class I and class II protein labels were randomly reassigned while preserving the original class sizes (172 class I and 195 class II proteins). For each of 1,000 permutations, we counted the number of significant within-class protein pairs at the corresponding locus and compared the observed count with the resulting permutation distribution. Permutation *P*-values were calculated as the proportion of permutations with a number of significant pairs equal to or greater than the observed value.

### Gene-level HLA colocalisation

To identify proteins whose HLA-pQTL signals share a common causal HLA gene with multiple sclerosis (MS) or Epstein–Barr virus (EBV) serological phenotypes, we applied *hlacoloc* (18), a Bayesian framework designed specifically for gene-level colocalisation for the classical HLA genes. Briefly, *hlacoloc* runs *SuSiE* (Sum of Single Effects regression) on HLA allele-level association statistics from each phenotype independently to compute per-allele posterior inclusion probabilities (PIPs), then performs Bayesian regression of the PIPs from the two phenotypes against each other to estimate the probability of a shared causal allele, reported as a per-gene colocalisation probability.

We included the 2-field classical HLA alleles for proteins with at least one significant HLA association. Where a significant association was identified at either 1-field or 2-field resolution for a given gene, all 2-field alleles for that protein were included in the analysis to provide full gene-level context for *SuSiE* fine-mapping. This resulted in 456 proteins taken forward for colocalisation testing.

Colocalisation was tested between joint analysis HLA-pQTL summary statistics and two disease phenotypes: MS risk from the International Multiple Sclerosis Genetics Consortium IMSGC (n = 47,850; 17,465 cases and 30,385 controls) and EBV serology including viral capsid antigen (VCA; n =7,741) and EBV nuclear antigen-1 (EBNA; n = 7,247). IgG titres measured in UK Biobank participants. For the pQTL data, we computed an in-sample LD matrix of classical HLA alleles at 2-field resolution using PLINK2 from the joint analysis genotype data. For the MS and EBV phenotypes, we used the external UK Biobank LD reference panel provided by Butler-Laporte et al. (18). This reference panel excluded UK Biobank participants with measured EBV antibody levels, ensuring full independence between the summary statistics from both phenotypes and the LD reference panel. The ‘is_cohort_ld’ flag was set to TRUE for the pQTL phenotype and FALSE for disease phenotypes.

Colocalisation was considered significant at a posterior probability threshold > 0.9 with concordant direction of effect. Results were reported separately for each classical HLA gene.

## Data availability

The UKB, CKB and joint pQTL summary statistics generated in this study will be deposited in Figshare upon publication.

## Code availability

Data and code to reproduce figures are available at: https://github.com/sarahsargg/mhc-pqtl-analysis

## Acknowledgements

We would like to acknowledge the UKB and CKB participants. All UKB data were accessed in accordance with UKB application number 31295 and 43920. This work was funded by a Kennedy Trust KTRR Senior Research Fellowship (KENN202109).

The CKB baseline survey and the first re-survey were supported by the Kadoorie Charitable Foundation in Hong Kong. Ongoing support was provided by the Wellcome Trust (212946/Z/18/Z, 202922/Z/16/Z, 104085/Z/14/Z, 088158/Z/09/Z), grants from the National Natural Science Foundation of China (82192901, 82192904, 82192900, 82388102), and the Noncommunicable Chronic Diseases-National Science and Technology Major Project (2023ZD0510101, 2023ZD0510100). DNA extraction and genotyping was funded by GlaxoSmithKline and the UK Medical Research Council (MC-PC-13049, MC-PC-14135). The project is supported by core funding from the UK Medical Research Council (MC_UU_00017/1, MC_UU_12026/2, MC_U137686851), Cancer Research UK (C16077/A29186; C500/A16896), and the British Heart Foundation (CH/1996001/9454) to the Clinical Trial Service and Epidemiological Unit and the MRC Population Health Research Unit at Oxford University.

This work was also supported by the Chinese Academy of Medical Sciences (CAMS) Innovation Fund for Medical Science (CIFMS), China (grant number: 2024-I2M-2-001-1), and a Nuffield Department of Medicine Career Development Award grant to AJM.

The computational aspects of this research were supported by the Wellcome Trust Core Award Grant Number 203141/Z/16/Z and the NIHR Oxford BRC. The views expressed are those of the author(s) and not necessarily those of the NHS, the NIHR or the Department of Health. GDS works within the MRC Integrative Epidemiology Unit at the University of Bristol, which is supported by the Medical Research Council (MC_UU_00032/01).

We are grateful to Krishna et al. for sharing their pQTL summary statistics.

## Author contributions

Conceptualization: SS, YL, SM, AM; Data curation: SS, SM, RN, YL, EN; Formal analysis: SS, SM, EN; Methodology: SS, YL, SM; Supervision: YL, SM, AM; Writing – original draft: SS, YL, SM; all authors contributed to the review and editing of the manuscript.

