## Supplementary Figures for "Multi-ancestry MHC-pQTL mapping reveals disease-linked HLA protein networks and shared genetic architecture"

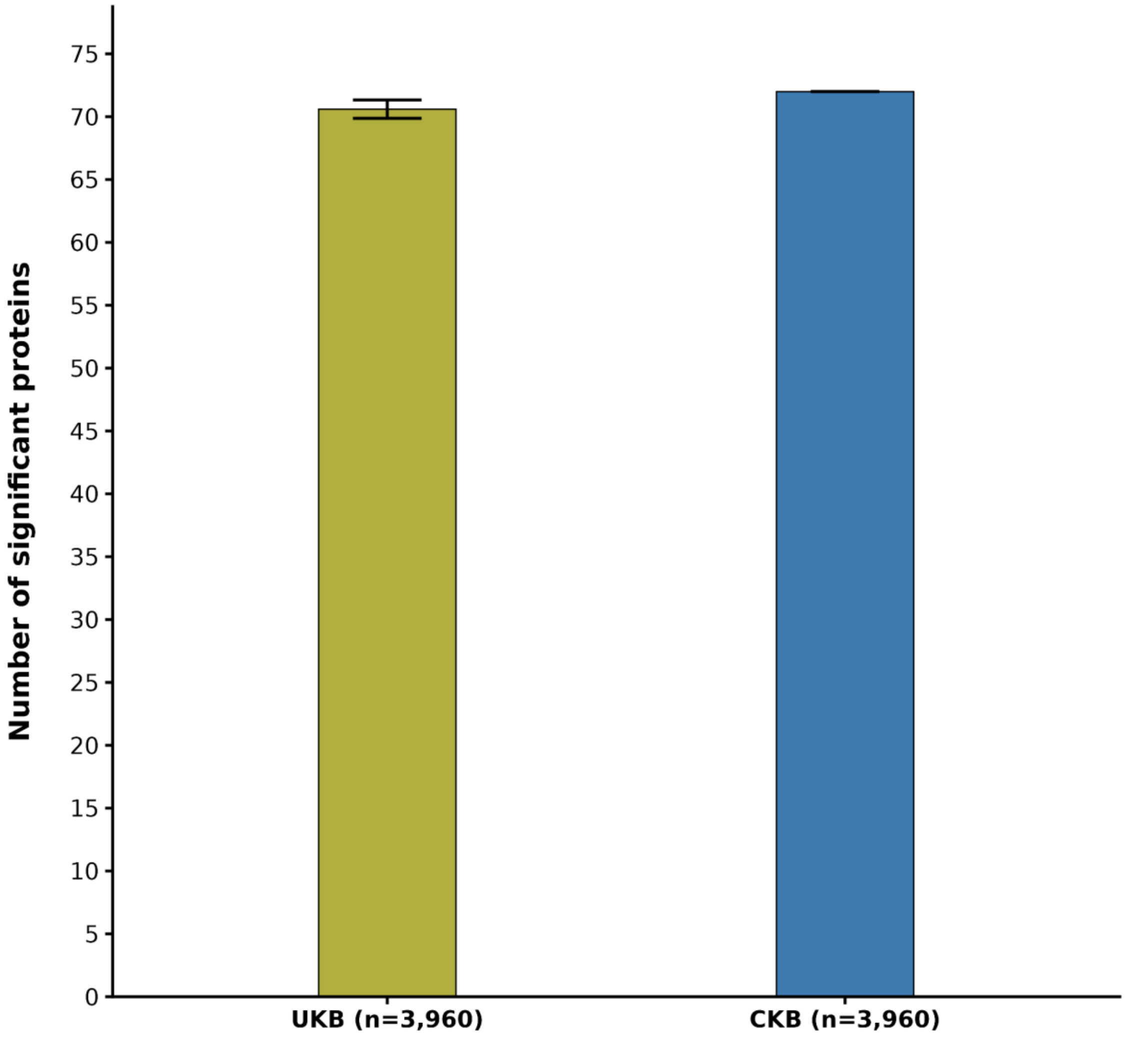


**Supplementary Figure 1. Comparable protein discovery between UKB and CKB at matched sample sizes.** Number of proteins with genome-wide significant MHC associations ($P < 1.71 \times 10^{-11}$) in downsampled UKB (n = 3,960; mean ± SE across 10 random iterations) and CKB (n = 3,960).

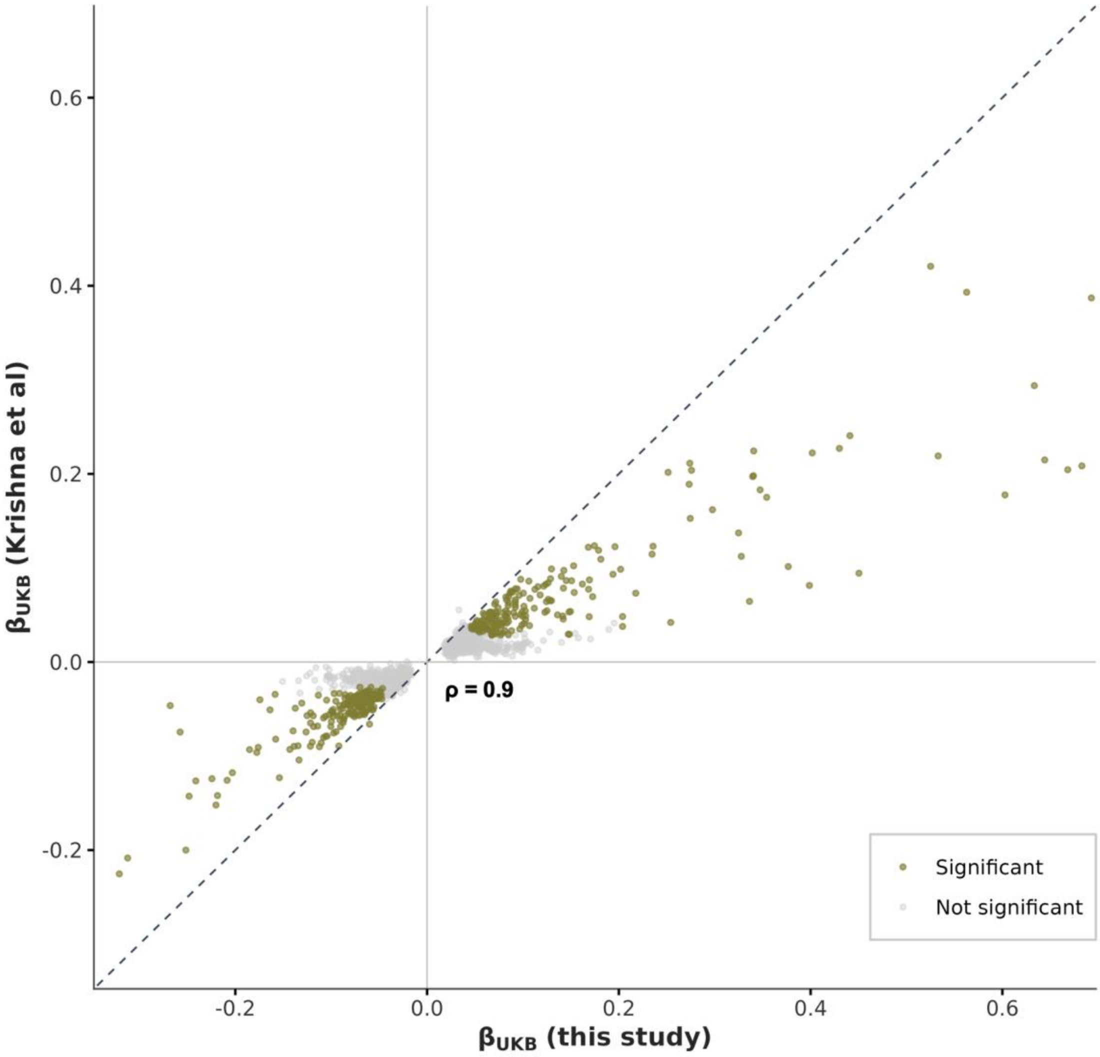


**Supplementary Figure 2. Replication of MHC-pQTL effect sizes against Krishna et al.** Effect size comparison of lead MHC-pQTL associations in UKB between this study (x-axis) and Krishna et al. discovery cohort (y-axis). Each point represents a protein-variant pair. The dashed line indicates perfect concordance. Strong agreement was observed across studies (Spearman ρ = 0.9).

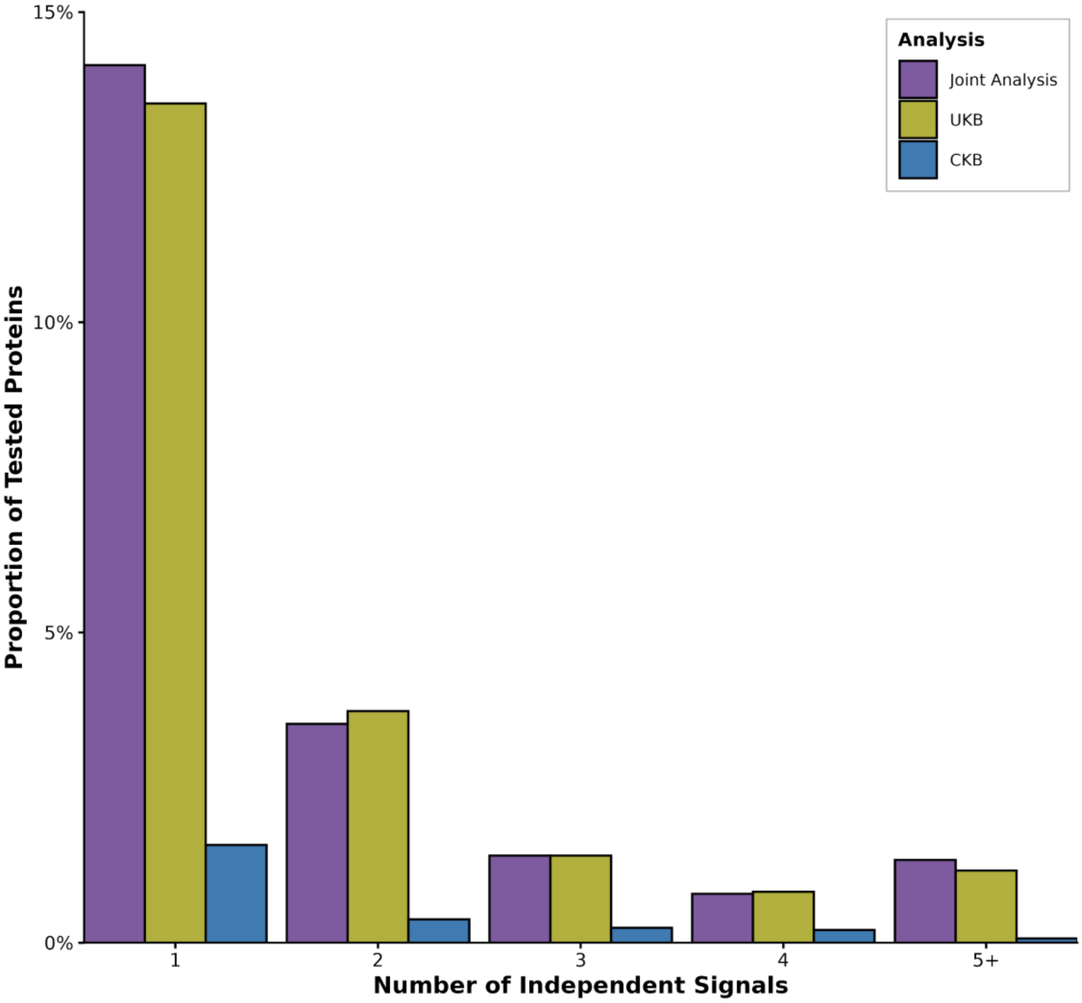


**Supplementary Figure 3. Distribution of independent association signals per protein across multi-ancestry analysis models.** Proportions are calculated relative to all tested proteins (n = 2,920). Independent signals were identified by iterative conditional analysis at a Bonferroni-corrected significance threshold ($P < 1.71 \times 10^{-11}$).

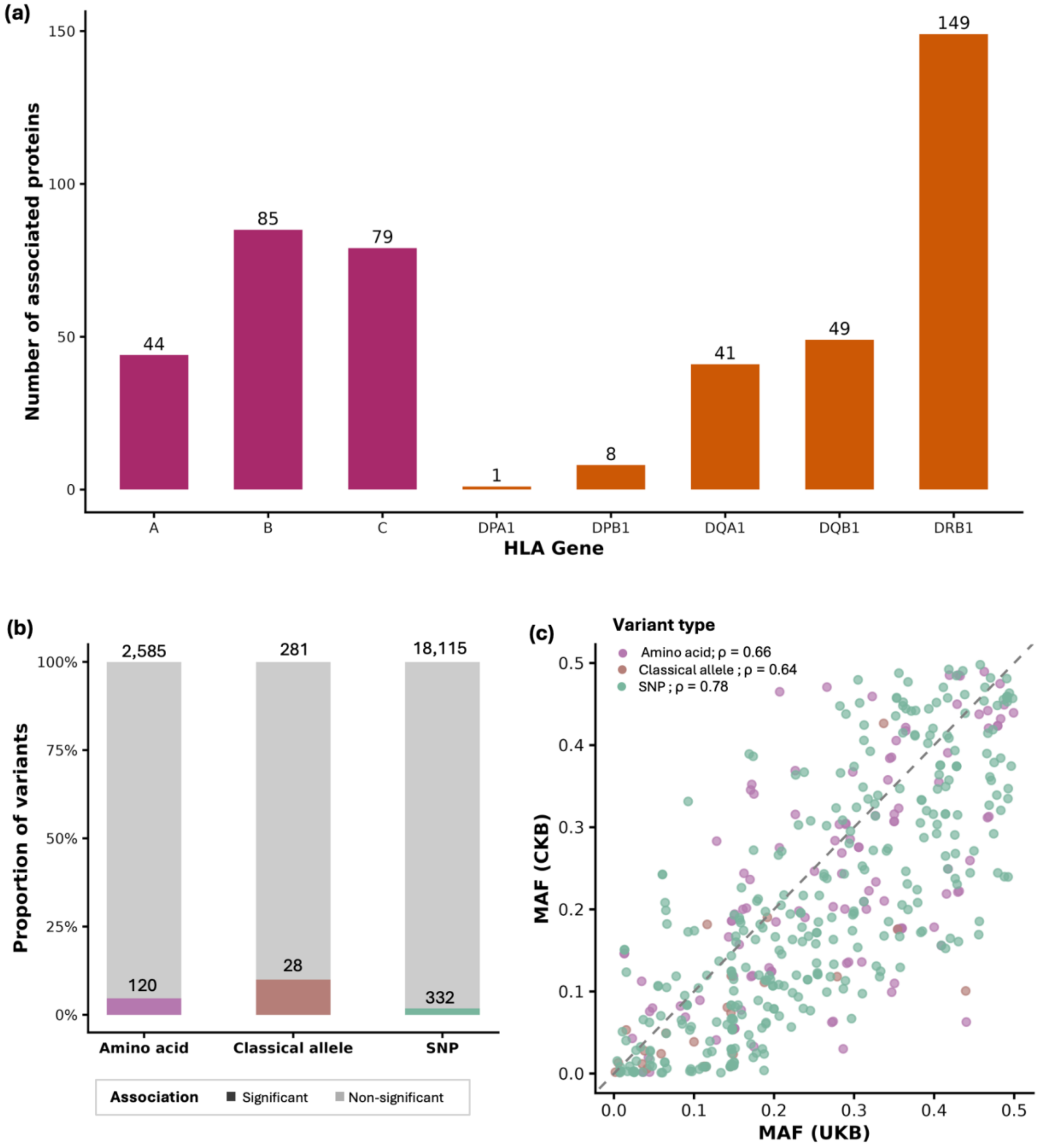


**Supplementary Figure 4. Genetic architecture of MHC-pQTL signals across HLA genes and variant types. (a)** Number of proteins with at least one significant classical HLA allele association ($P < 1.71 \times 10^{-11}$) at each of the eight classical HLA genes. **(b)** Proportion of tested variants that were significant lead pQTL variants, stratified by variant type. Numbers within bars indicate the count of significant lead variants; numbers above bars indicate the total number of tested variants in each category. **(c)** Minor allele frequency comparison of lead pQTL variants between UKB (European ancestry) and CKB (East Asian ancestry), coloured by variant type. Spearman correlations are shown per variant type.

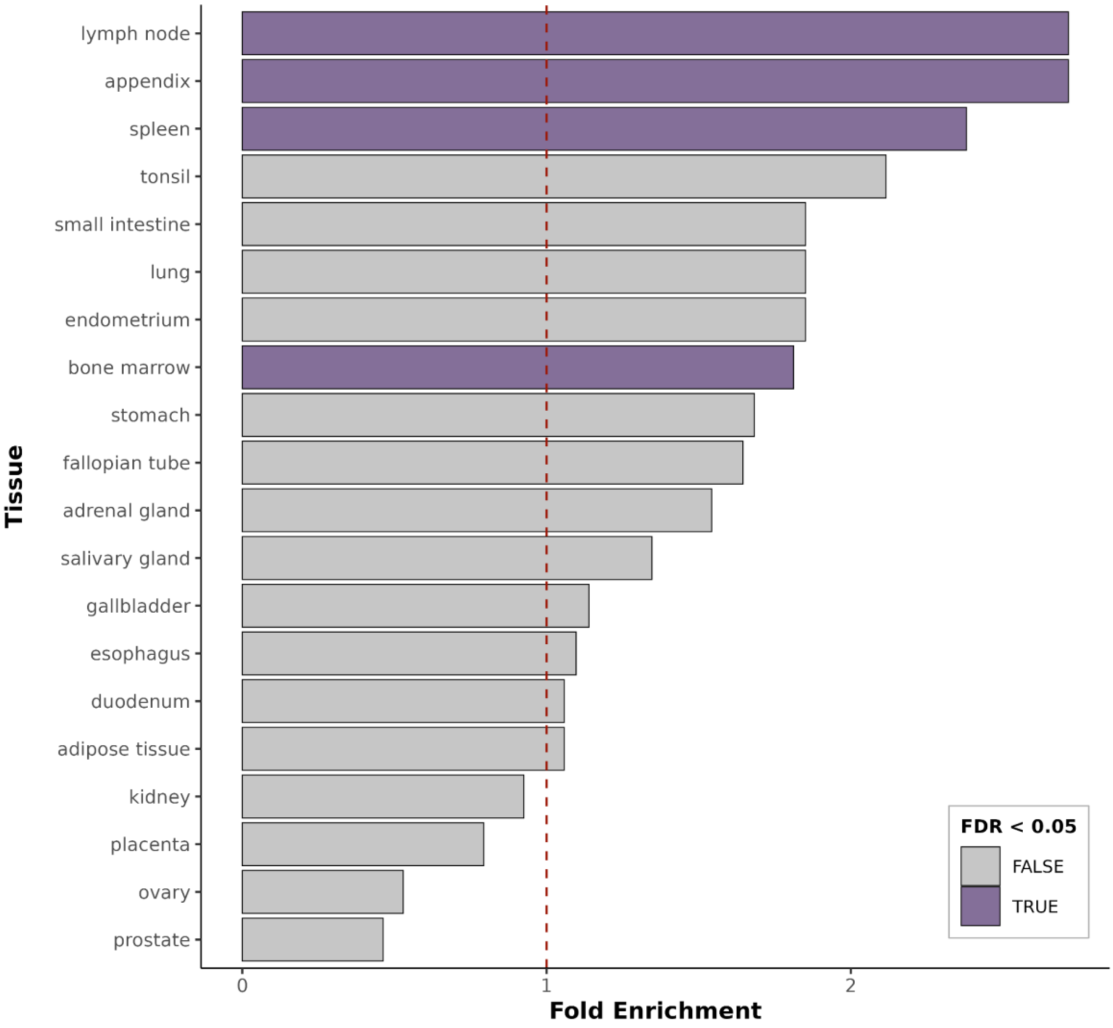


**Supplementary Figure 5. Tissue enrichment of MHC-regulated proteins.** Dashed line indicates fold enrichment of 1 (no enrichment). Bars are coloured by statistical significance after FDR correction.

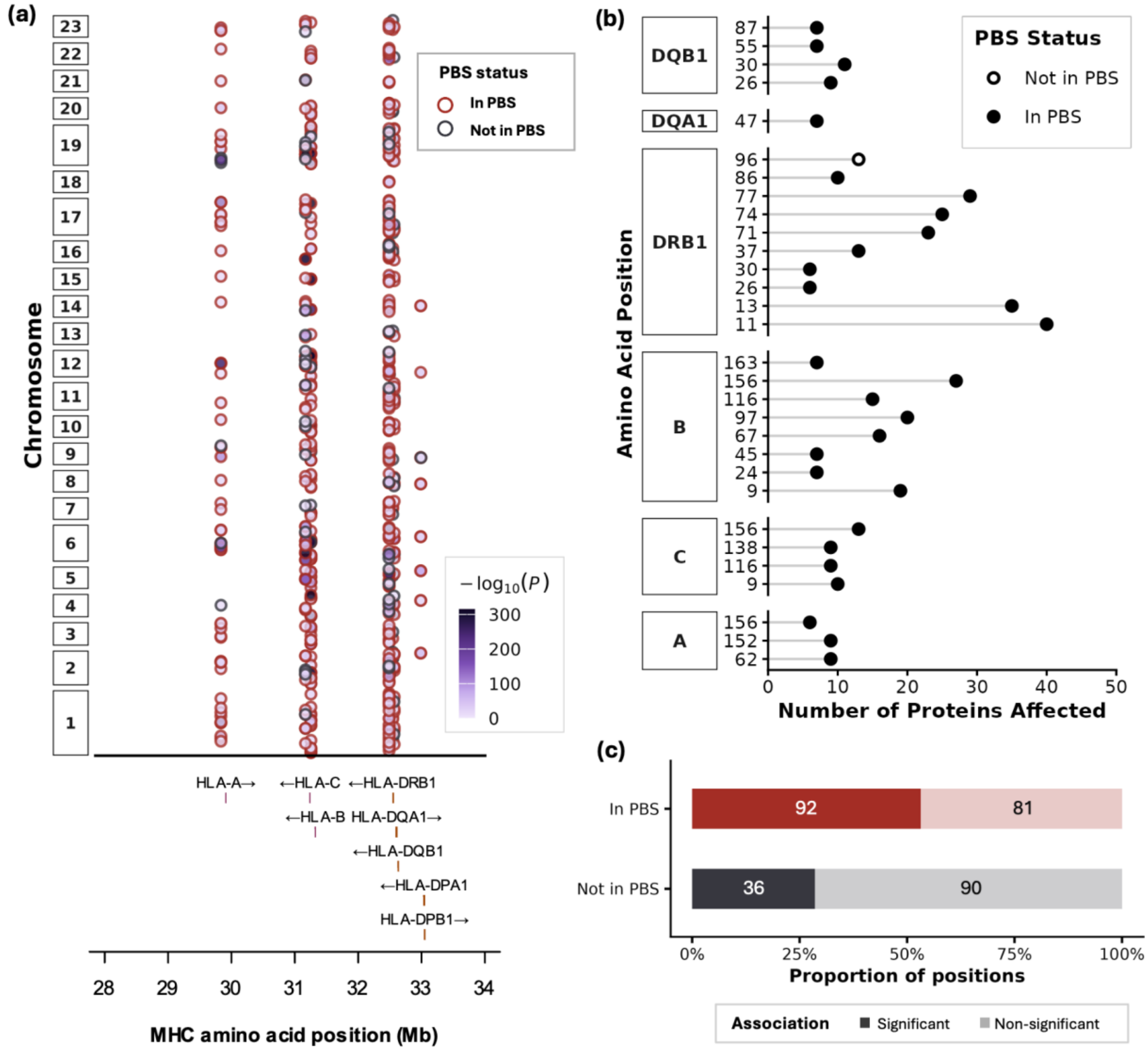


**Supplementary Figure 6. Amino acid fine-mapping of MHC-pQTL signals.** (a) Genomic distribution of significant protein-amino acid position associations from omnibus testing across the MHC region. Each point represents a significant association ($P < 1.71 \times 10^{-11}$), coloured by $-\log_{10}(P)$. Points are outlined in red if the amino acid position falls within the peptide-binding groove (defined by exons 2 and 3 for HLA class I genes and exon 2 for class II genes) and in grey otherwise. (b) Number of proteins significantly associated with each amino acid position, grouped by HLA gene. Only positions associated with more than 5 proteins are shown. (c) Proportion of amino acid positions with at least one significant protein association, stratified by location within or outside the peptide-binding groove.

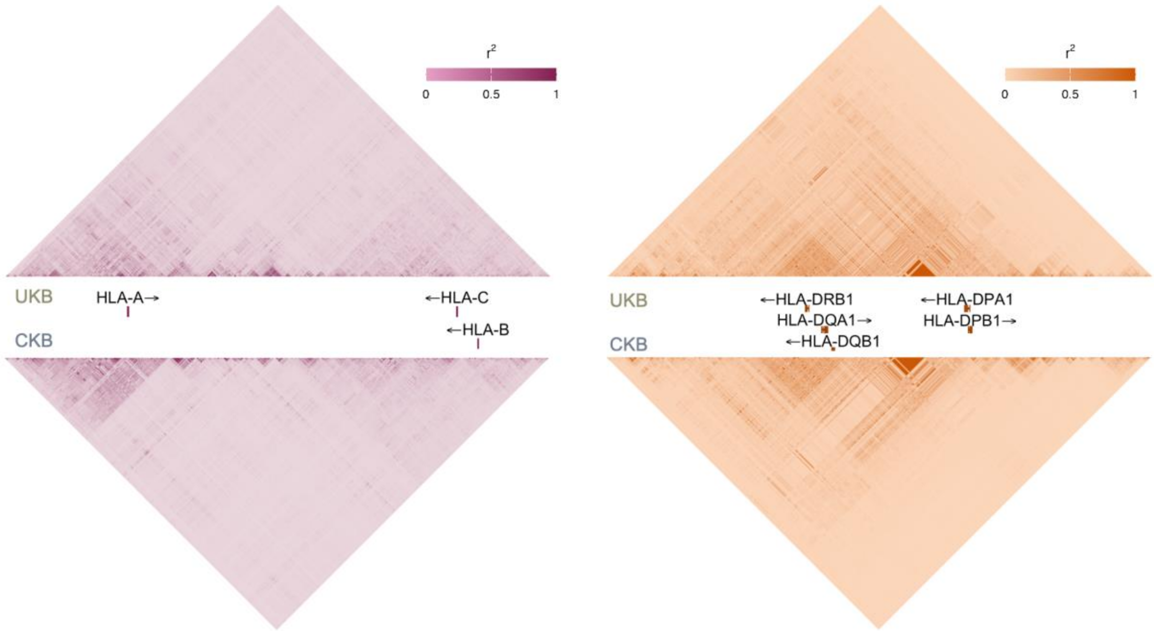


**Supplementary Figure 7. Linkage disequilibrium structure within HLA class I and class II regions across UKB and CKB cohorts.** Pairwise LD ($r^2$) heatmaps for variants within the HLA class I region (left, HLA-A, -B, -C) and HLA class II region (right, HLA-DRB1, -DQA1, -DQB1, -DPA1, -DPB1).

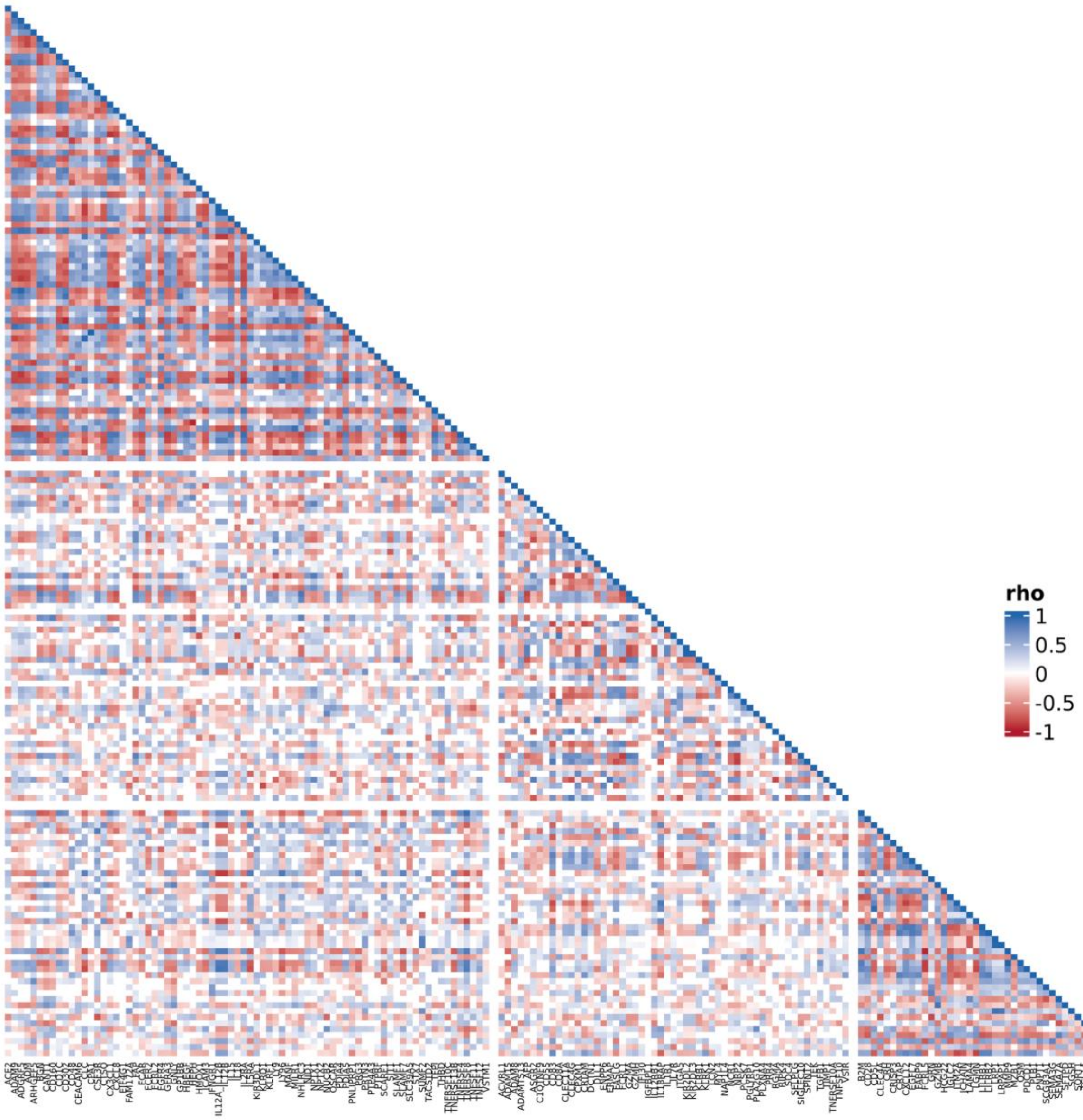


**Supplementary Figure 8. Local genetic correlation matrix among HLA class I-associated trans-pQTL proteins.** Pairwise local genetic correlations (ρ) estimated by LAVA within the HLA class I locus (chr6:29.5–31.5 Mb) for 172 proteins with class I-specific trans-pQTL associations. Blue indicates positive genetic correlation and red indicates negative genetic correlation, with colour intensity proportional to |ρ|. Proteins are ordered by Louvain community detection clustering.

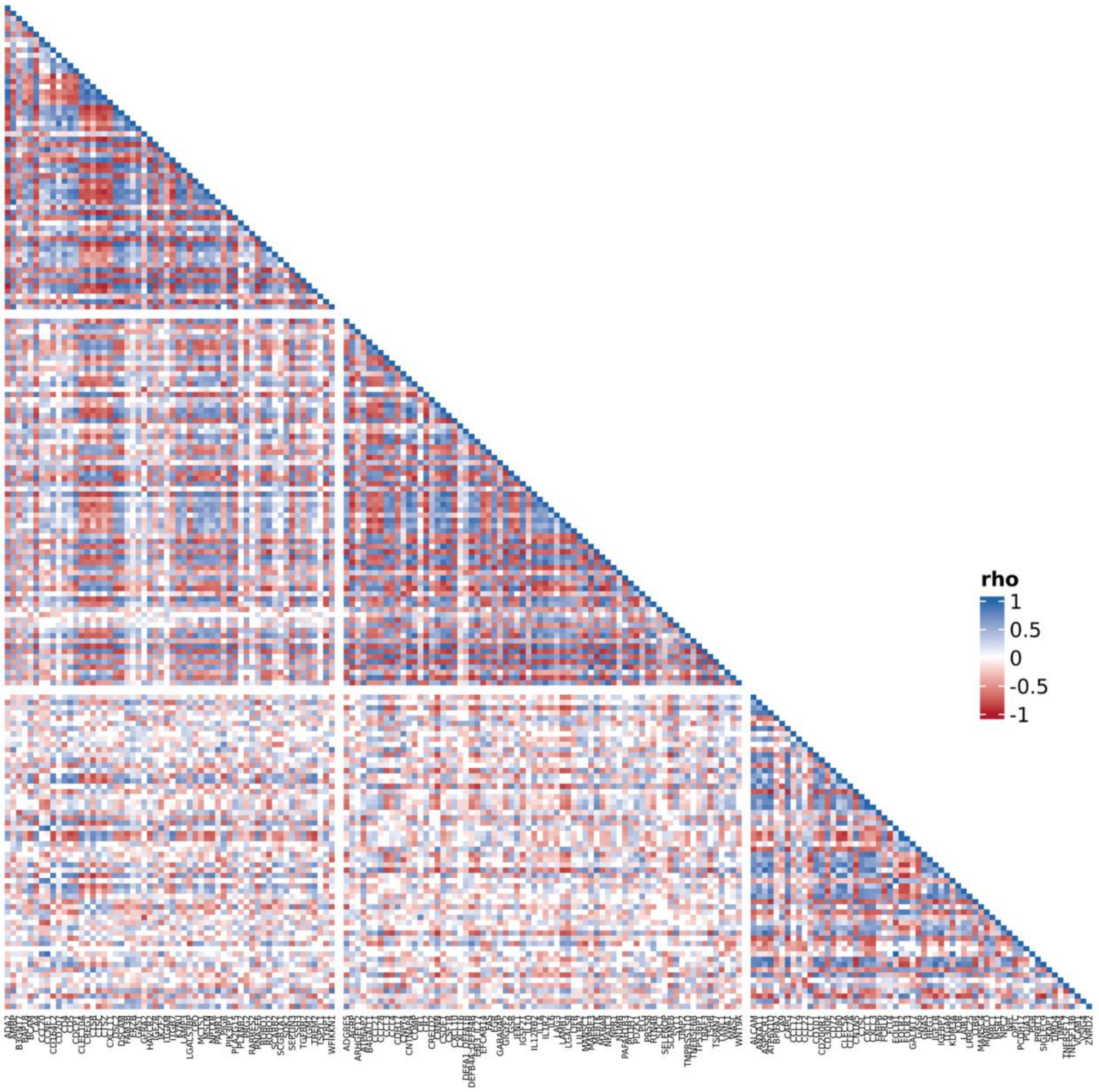


**Supplementary Figure 9. Local genetic correlation matrix among HLA class II-associated trans-pQTL proteins.** Pairwise local genetic correlations (ρ) estimated by LAVA within the HLA class II locus (chr6:32.0–33.5 Mb) for 195 proteins with class II-specific trans-pQTL associations. Blue indicates positive genetic correlation and red indicates negative genetic correlation, with colour intensity proportional to |ρ|. Proteins are ordered by Louvain community detection clustering.

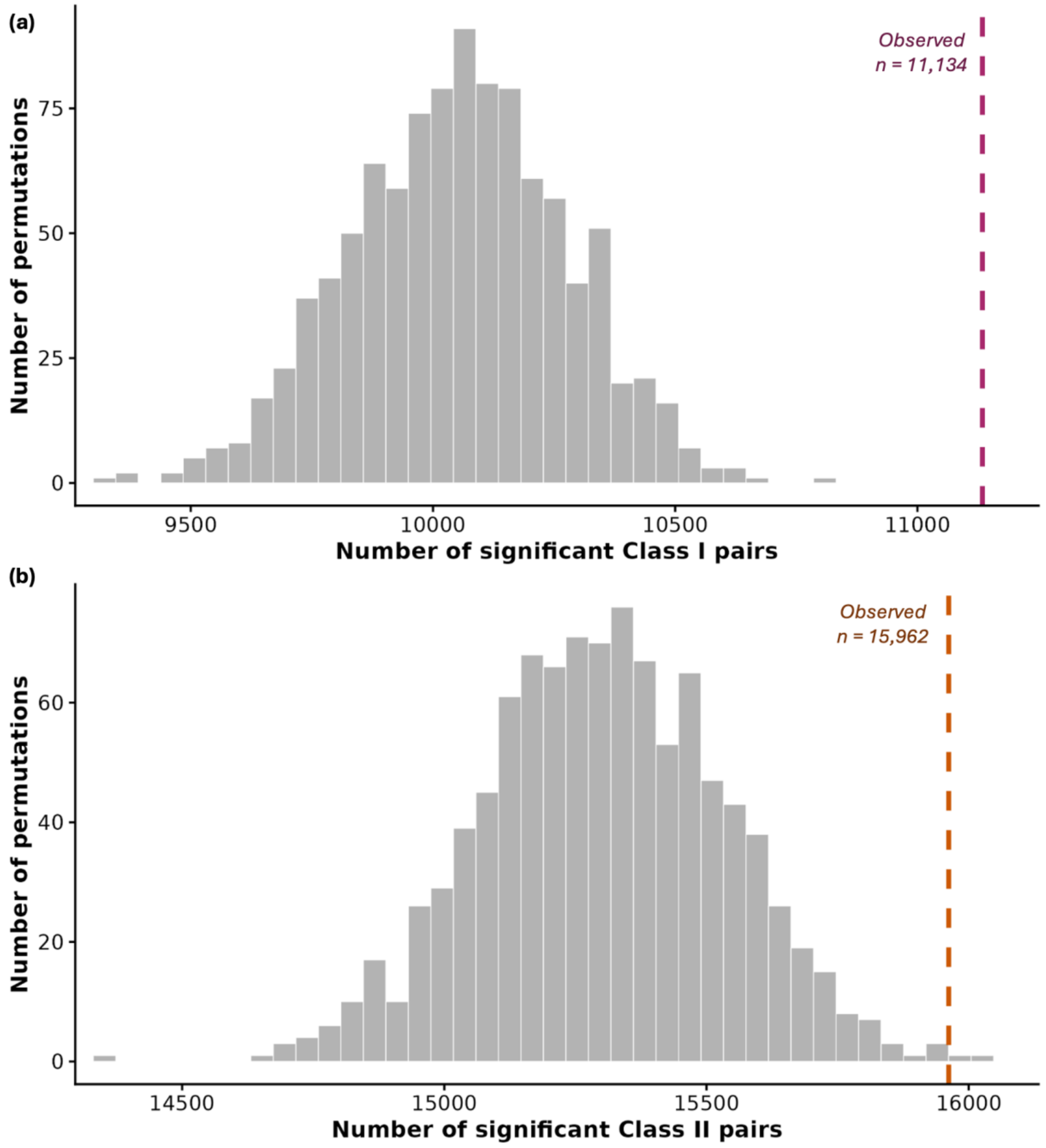


**Supplementary Figure 10.** Permutation analysis of within-class significant protein pairs. Distribution of the number of significant within-class protein pairs obtained across 1,000 random permutations of HLA class assignments while preserving the original class sizes. (a) Class I locus, showing the observed number of significant Class I–Class I pairs (11,134; dashed line). (b) Class II locus, showing the observed number of significant Class II–Class II pairs (15,962; dashed line). The observed values fall beyond the permutation distributions, supporting non-random enrichment of within-class significant local genetic correlations.

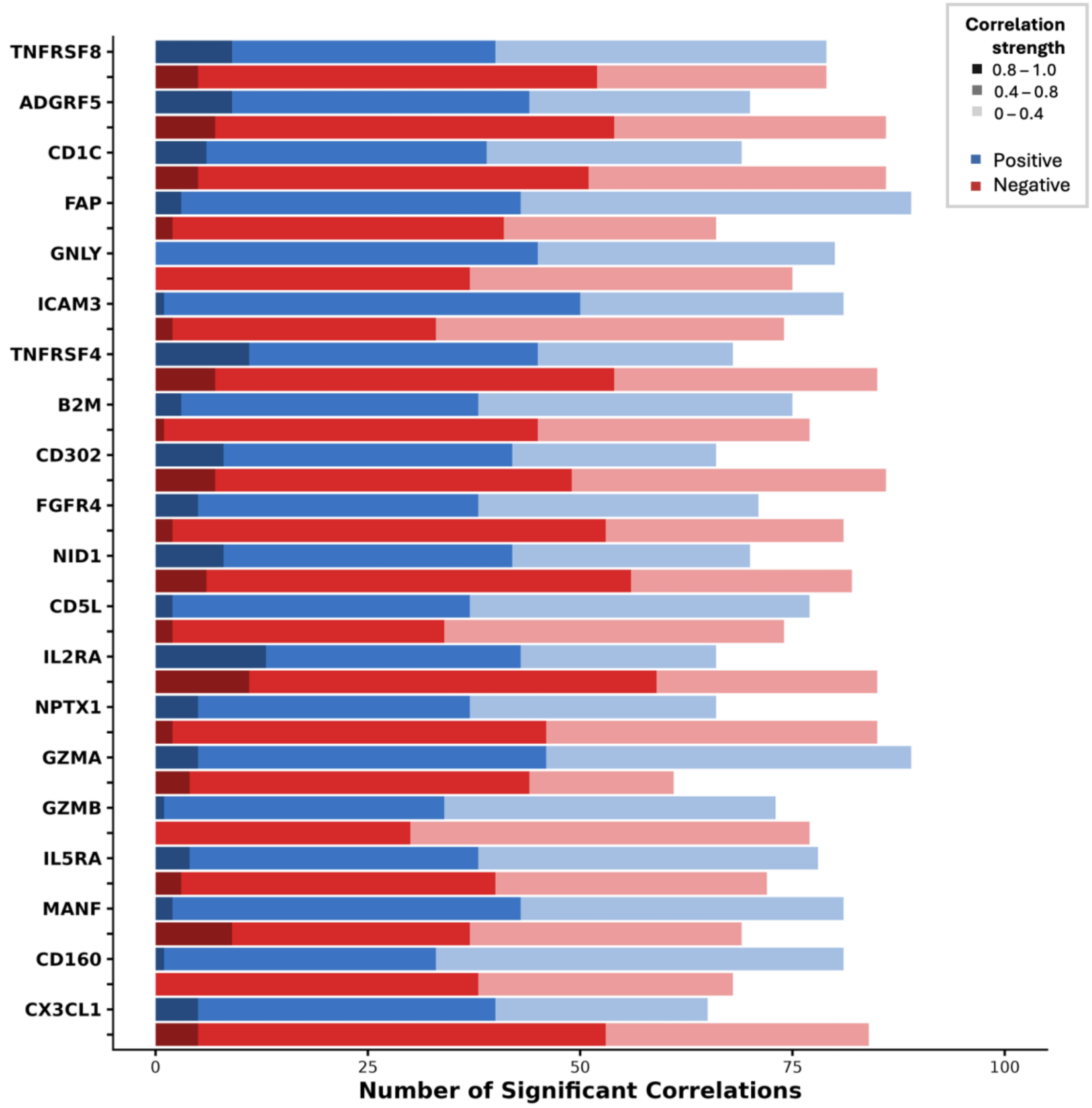


**Supplementary Figure 11. Correlation strength and connectivity of the top 20 most connected proteins in the LAVA co-regulation networks in HLA class I.**
Positive correlations are shown in blue and negative correlations in red. Bar length indicates the number of significant pairwise correlations, and colour intensity reflects correlation strength, with darker shades indicating stronger correlations. Significant correlations were defined as $P < 1.55 \times 10^{-6}$.

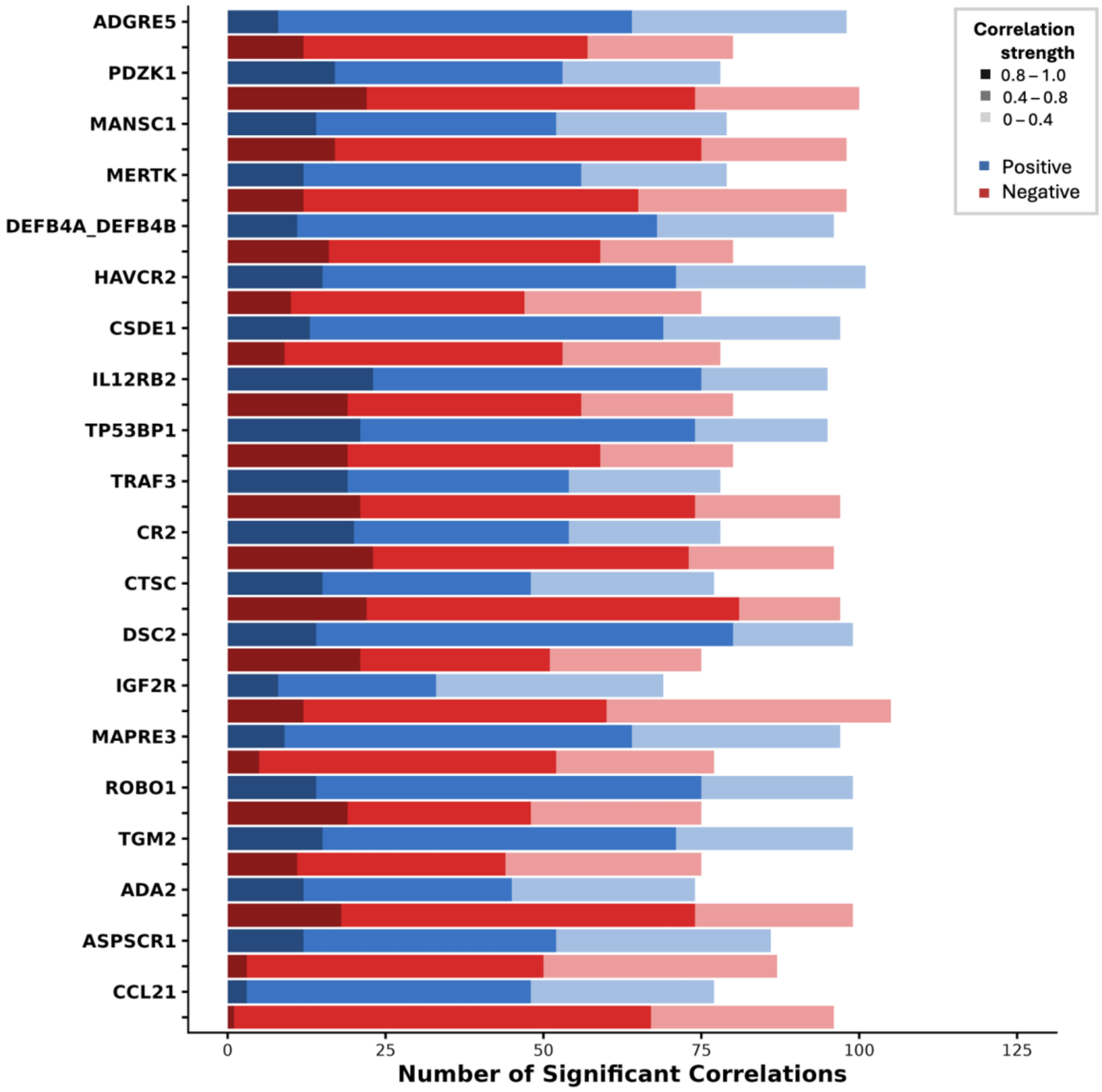


**Supplementary Figure 12. Correlation strength and connectivity of the top 20 most connected proteins in the LAVA co-regulation networks in HLA class II.**

Positive correlations are shown in blue and negative correlations in red. Bar length indicates the number of significant pairwise correlations, and colour intensity reflects correlation strength, with darker shades indicating stronger correlations. Significant correlations were defined as $P < 1.55 \times 10^{-6}$.

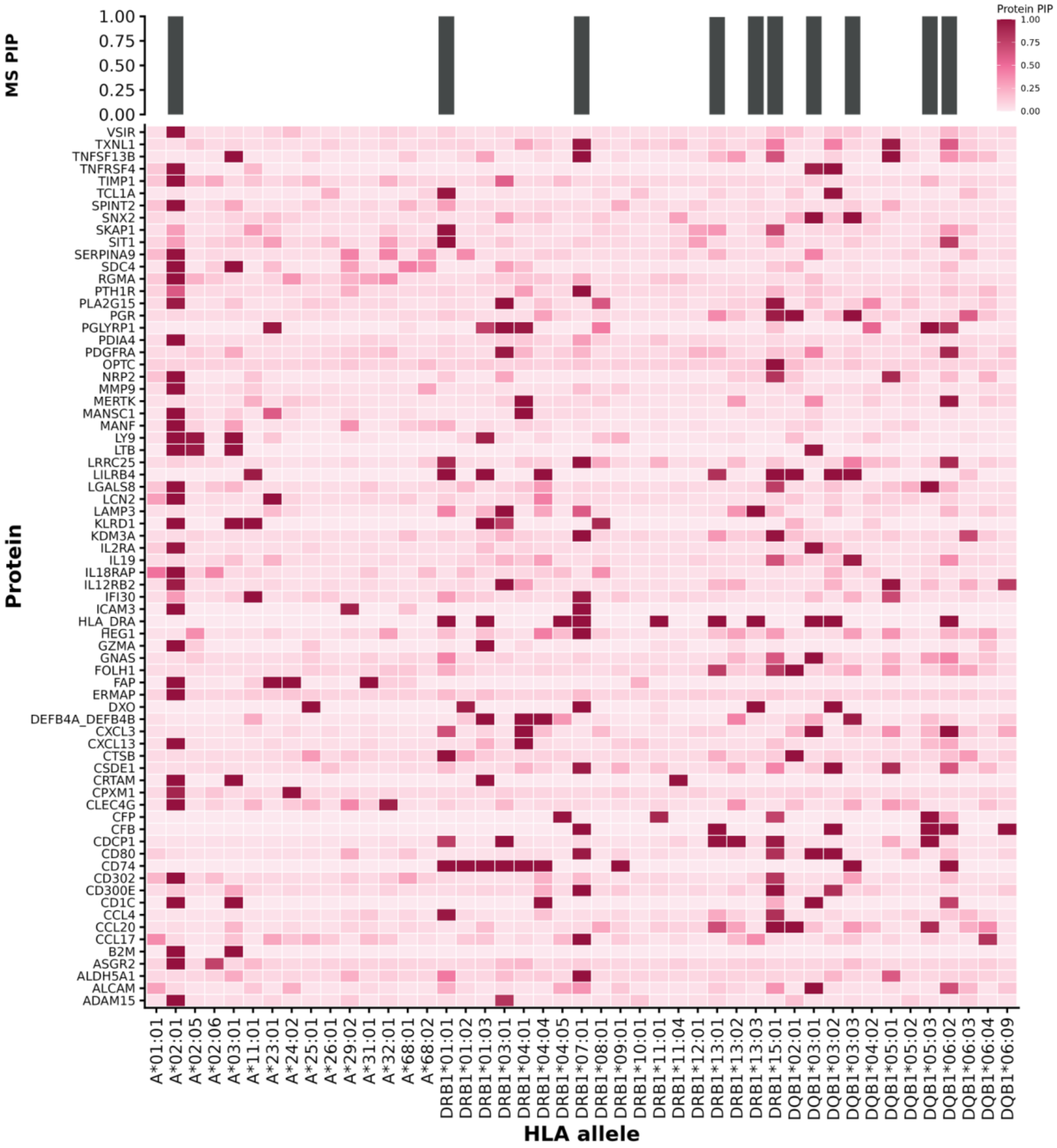


**Supplementary Figure 13. Posterior inclusion probability profiles of MS-colocalised proteins across HLA alleles.** Heatmap of per-allele posterior inclusion probabilities (PIPs) from SuSiE fine-mapping for proteins colocalising with multiple sclerosis (posterior probability > 0.90). Rows represent colocalised proteins; columns represent classical HLA alleles at HLA-A, HLA-DRB1 and HLA-DQB1. Cell colour intensity indicates the protein-side PIP, with darker cells denoting higher probability that the allele is causal for the protein's pQTL signal. The top bar chart shows the corresponding MS-side PIPs per allele. Notable convergence is visible at HLA-A*02:01 for class I-regulated proteins.

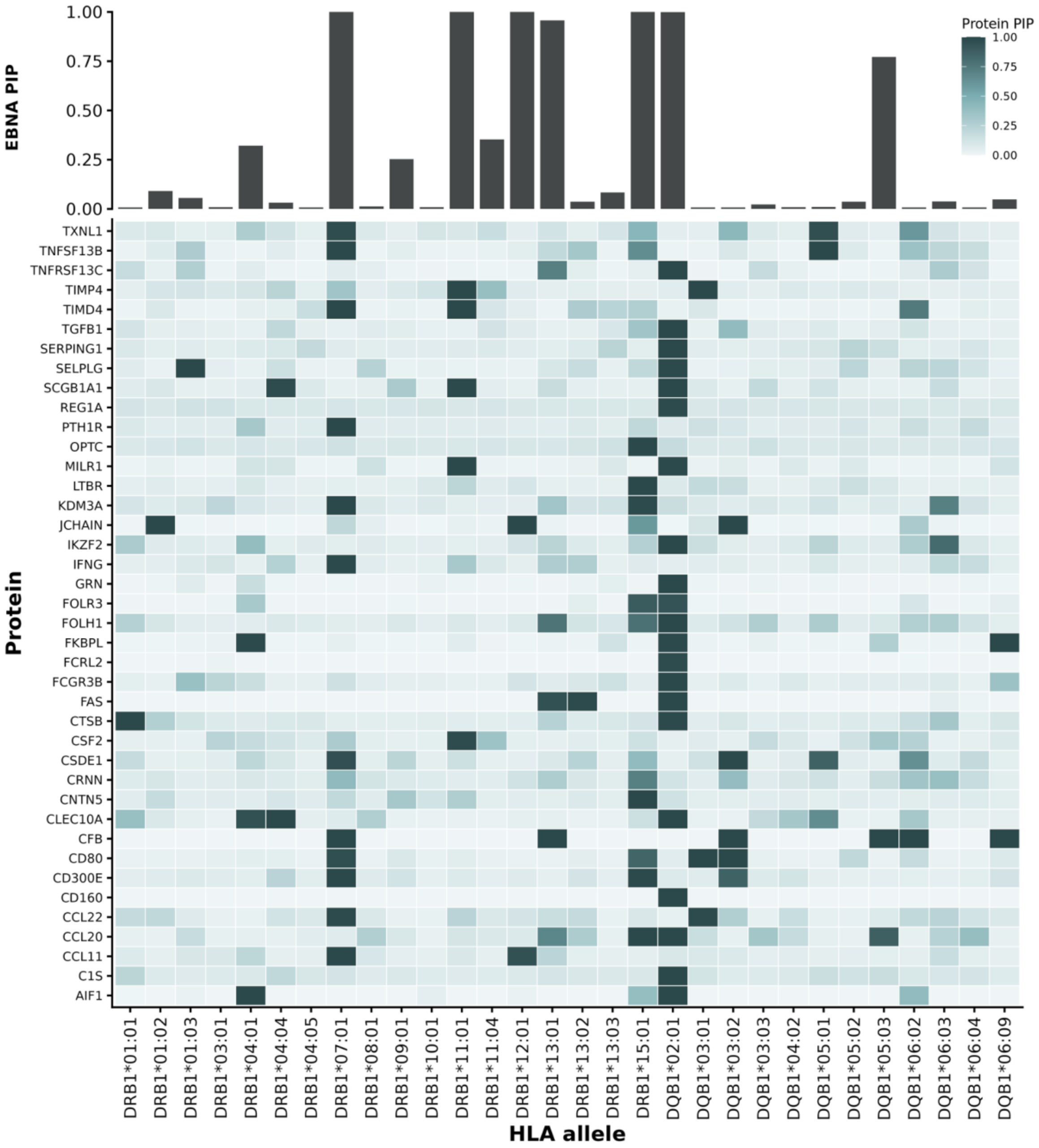


**Supplementary Figure 14. Posterior inclusion probability profiles of EBNA-colocalised proteins across HLA alleles.** Heatmap of per-allele posterior inclusion probabilities (PIPs) from SuSiE fine-mapping for proteins colocalising with EBNA antibody response (posterior probability > 0.90). Rows represent colocalised proteins; columns represent classical HLA alleles at HLA-DRB1 and HLA-DQB1. Cell colour intensity indicates the protein-side PIP. The top bar chart shows the corresponding EBNA-side PIPs per allele.

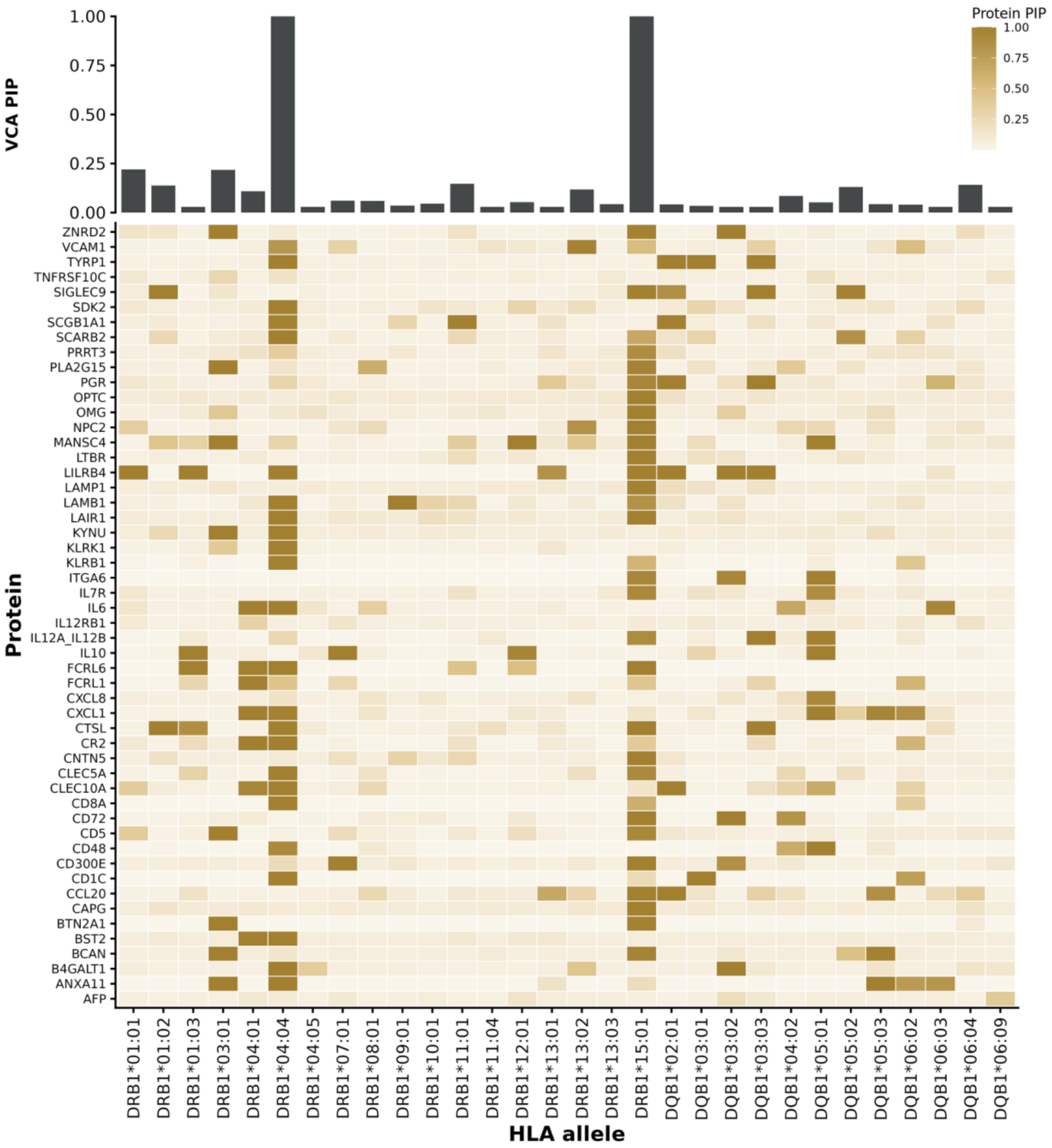


**Supplementary Figure 15. Posterior inclusion probability profiles of VCA-colocalised proteins across HLA alleles.** Heatmap of per-allele posterior inclusion probabilities (PIPs) from SuSiE fine-mapping for proteins colocalising with VCA antibody response (posterior probability > 0.90). Rows represent colocalised proteins; columns represent classical HLA alleles at HLA-DRB1 and HLA-DQB1. Cell colour intensity indicates the protein-side PIP. The top bar chart shows the corresponding VCA-side PIPs per allele. VCA signal is concentrated at DRB1*04:05 and DRB1*15:01, with broad protein PIP enrichment at these alleles consistent with HLA control of anti-lytic EBV antibody responses through class II-restricted immune pathways.

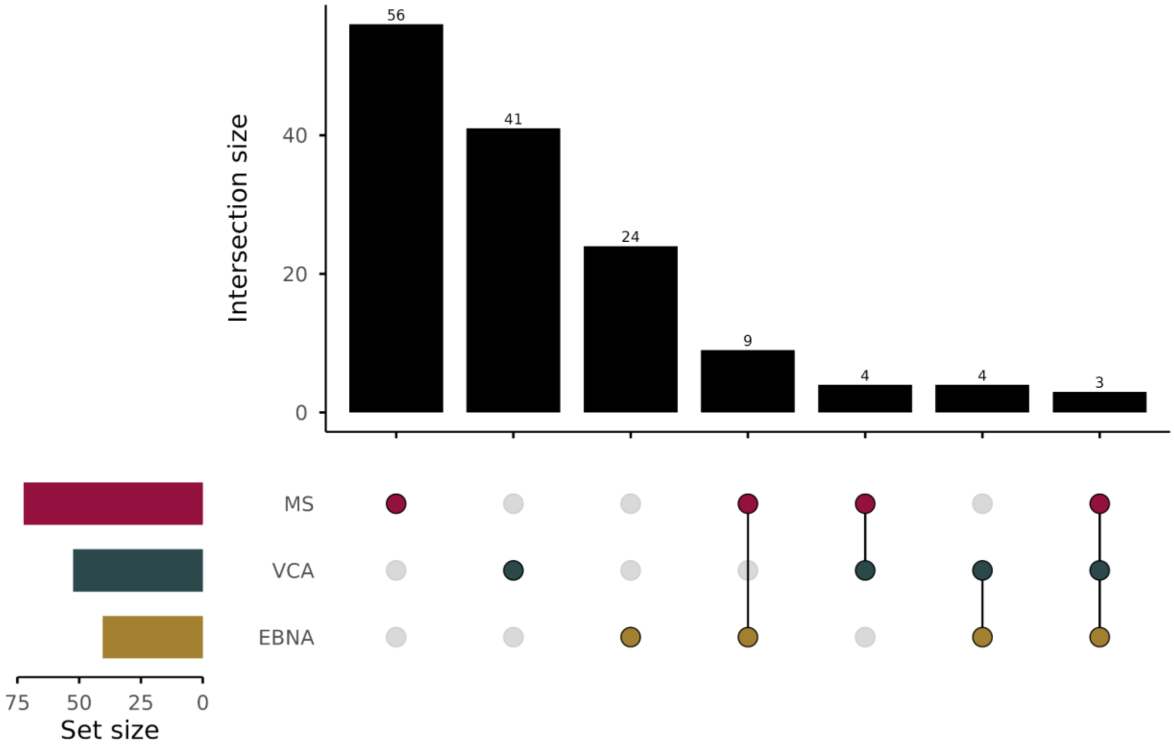


**Supplementary Figure 16. Overlap of colocalised proteins across MS and EBV serological endpoints.** UpSet plot showing the intersection of proteins with significant HLA colocalisation (posterior probability > 0.90) across multiple sclerosis (MS), viral capsid antigen (VCA) antibody response and Epstein-Barr nuclear antigen (EBNA) antibody response.

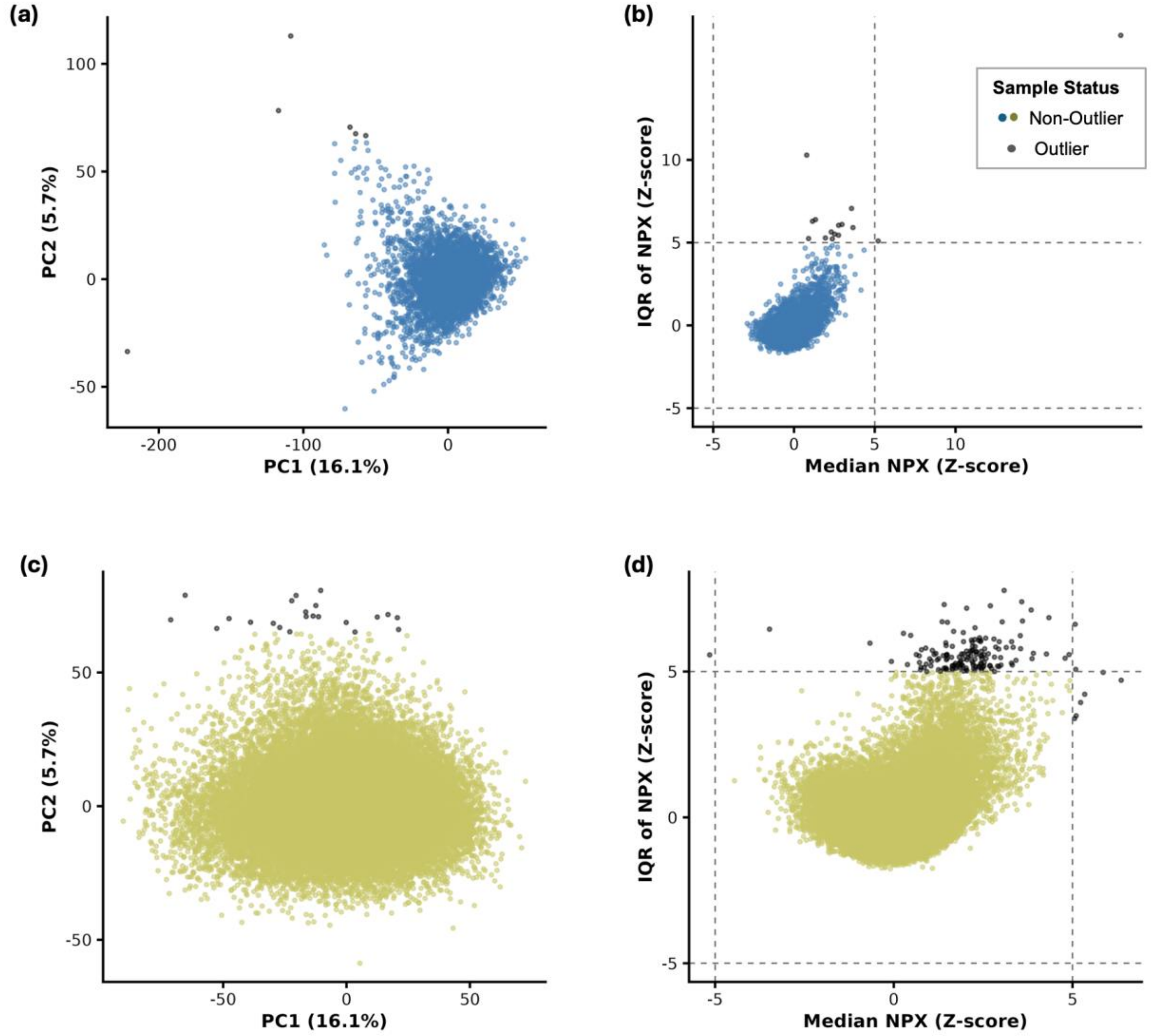


**Supplementary Figure 17. Sample-level outlier detection in proteomic data.** Outlier identification in CKB (a, b) and UKB (c, d) using two complementary approaches. (a, c) PCA of the imputed proteomic matrix, showing PC1 versus PC2. Individuals exceeding ± 5 standard deviations on either component were flagged as outliers. (b, d) Per-sample median NPX (Z-scored) versus IQR of NPX (Z-scored). Dashed lines indicate the ± 5 standard deviation thresholds; individuals exceeding these thresholds on either metric were flagged. The union of outliers from both methods was removed prior to downstream analysis (UKB: 180 removed; CKB: 17 removed).

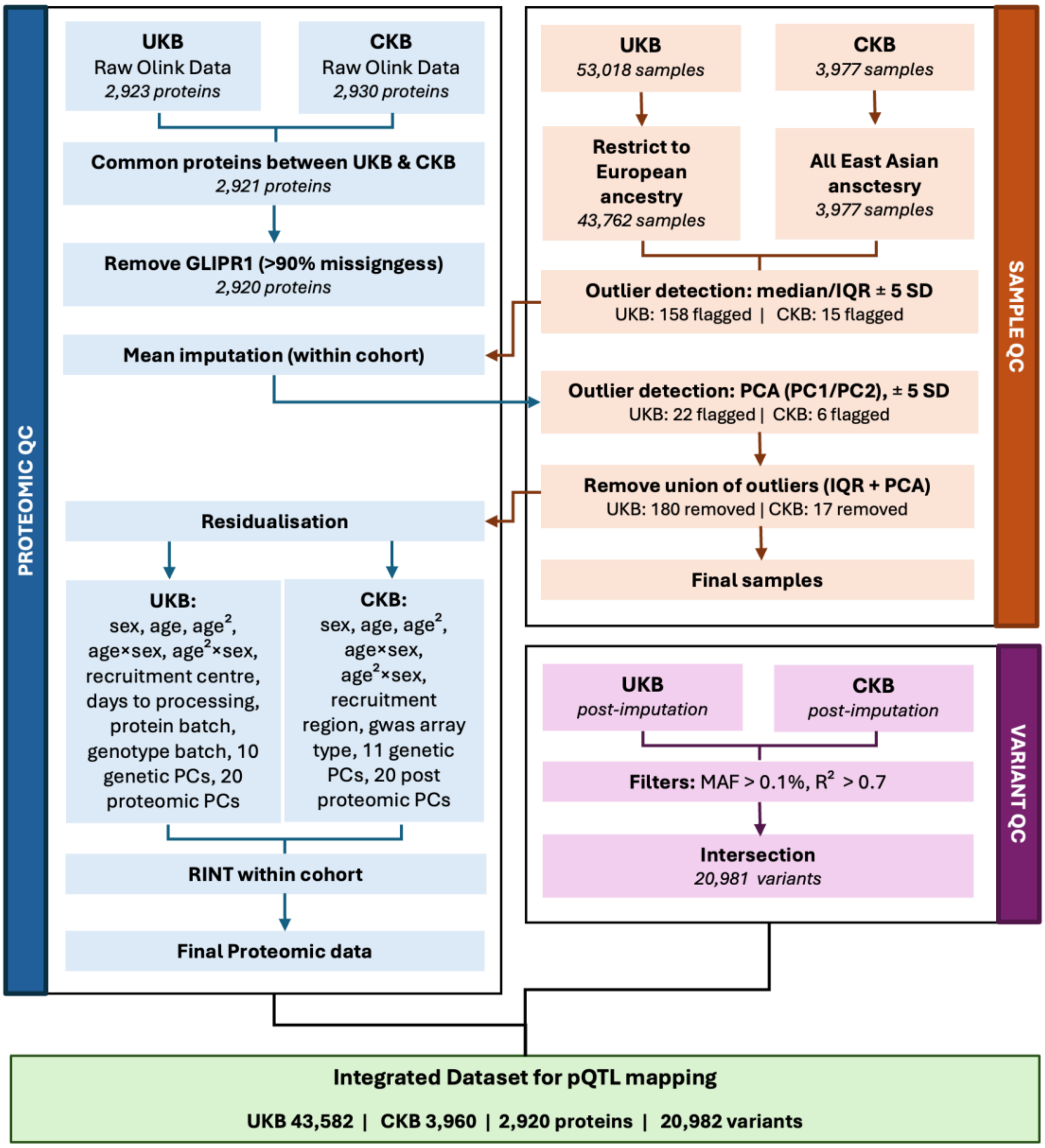


**Supplementary Figure 18. Quality control and data integration pipeline.** Overview of sample, protein and variant quality control steps applied to UKB and CKB proteomic and genotype data. The final integrated dataset comprised 43,582 UKB and 3,960 CKB individuals across 2,920 proteins and 20,981 MHC-region variants.

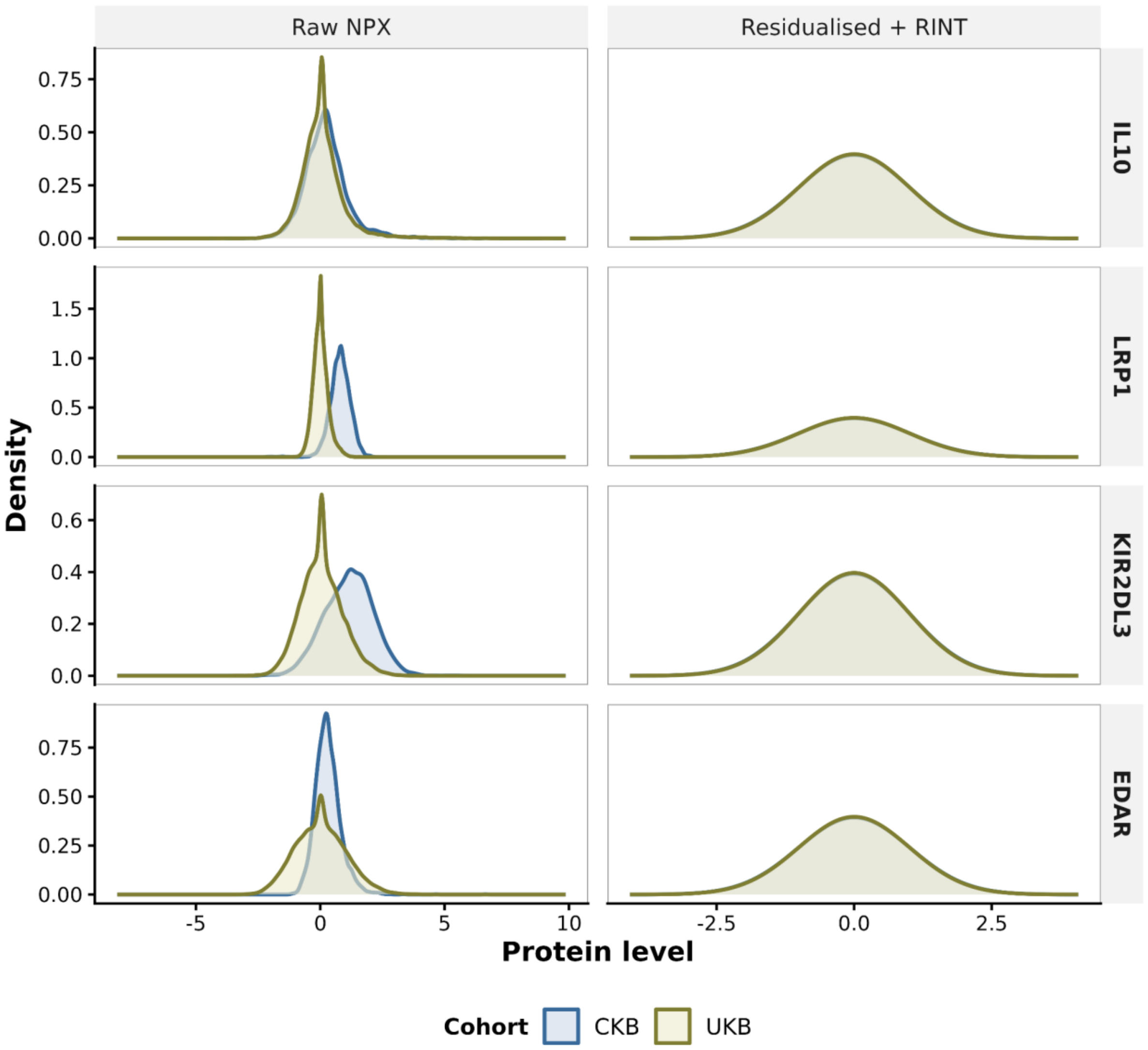


**Supplementary Figure 19. Pre- and post-normalisation protein expression distributions.** Density plots of raw NPX values (left) and residualised + RINT-transformed values (right) for four representative proteins, stratified by cohort.
